# Pitfalls in understanding how multiple long-term conditions cluster: whole population and age-stratified associations in 7,490,874 people in England

**DOI:** 10.1101/2025.02.06.25321779

**Authors:** Guillermo Romero Moreno, Valerio Restocchi, Nazir Lone, Jacques D. Fleuriot, Jake Palmer, Luna De Ferrari, Bruce Guthrie

## Abstract

Studies of how multiple long-term conditions (MLTC) cluster together in individuals vary in the populations studied, and whether they age and/or sex stratify, which limits comparison between studies and reproducibility. This study uses a large, UK primary-care dataset to examine how pairwise strength of association between 74 conditions varies by age in both men and women aged 30-99 years, and to explore implications for MLT cluster analyses. Joint prevalence of conditions was lowest in younger age-groups and progressively increased with age, whereas Association Beyond Chance (ABC) was highest in younger age-groups and progressively decreased with age. Condition clustering based on ABC identified different clusters in all men and all women aged 30-99 years, and these clusters differed from those identified in each age-group. Researchers examining how MLTC cluster should consider whether age and sex stratification is appropriate given their study aims and/or would improve comparability and reproducibility, and explicitly justify their choices.

## Introduction

Population ageing and improved survival from acute life-threatening conditions is driving increasing prevalence of multimorbidity, i.e. the presence of multiple long-term conditions (MLTC) in the same individual.^27,36^ Having MLTC is associated with higher mortality, lower quality of life, and higher use of health and social care.^28,29^ Population ageing and increasing MLTC prevalence presents major challenges to health systems internationally, which are largely designed around care for single diseases. The same focus on single diseases also dominates health research, but there is increasing recognition that research into how MLTC develop, and their consequences, is important to address the challenges that MLTC pose.^36^

A better understanding of how long-term conditions (LTC) cluster, both in cross-sectional data and in terms of trajectories of individual morbidity accrual over time, has been identified as a priority for MLTC research by the UK Academy of Medical Sciences.^1^ Numerous studies examining how conditions are associated with each other have now been published, with a shift over time from methods like exploratory factor analysis towards network methods. However, systematic reviews of this literature identify problems with reproducibility and generalizability due to heterogeneity in choice of LTCs to include, measures of association or distance used, and analytical approaches.^2–4^ Variation is to be expected, since optimal choices will depend on the purpose of the analysis, where purposes can include both aetiological studies intended to inform understanding of common causes and mechanisms, and more applied studies intended to inform organization of care.

In addition, since we know that conditions typically have different ages of onset and vary in prevalence between sexes, it would be expected that age and sex will affect individual condition prevalence, associations between conditions, as well as the consequent patterns of clustering of conditions observed in the data. However, fewer than 10% of studies in a recent systematic review stratified their analysis by either age or sex, so clustering solutions have historically largely been applied to the whole population studied.^2^ Studies that did undertake stratification found that ‘cardiometabolic’ clusters were consistently observed across age and sex strata, but other clusters varied. For example, Prados-Torres observed a “mechanical-obesity-thyroidal” cluster in women of all ages, but only in men aged 45+, and also a “psychiatric-substance misuse” cluster which was only observed only in younger men.^7–10^ Busija et al’s 2019 systematic review included 51 LTCs clustering studies of which only 29% used stratified analyses in any way (at least in part because many studies were small),^15^ and concluded that although observed clusters were relatively stable in sex and ethnicity strata, there was more variation in cluster solutions observed in different age strata.

Variation in the age range and structure of the population studied is therefore likely to alter observed associations between LTCs and subsequent clusters, and likely contributes to the inconsistencies in findings in the published literature.^10^ The aim of this study was to systematically examine variation in pairwise associations between, and joint prevalence of, MLTC in analysis of a large population database stratified by age and sex, and explore the implications for clustering analysis of MLTC.

## Methods

### Data source and study population

We used a UK dataset of primary care Electronic Health Records from the Clinical Practice Research Datalink (CPRD),^39^ which provides rich information about the conditions patients have as well as demographic information. For our study, we selected patients permanently registered on 1^st^ January 2018 with a primary care general practice (GP) in England contributing to the CPRD Aurum dataset, and with linkage to hospital admission data (Health Episode Statistics – Admitted Patient Care [HES-APC]) and Office of National Statistics (ONS) mortality registration. Eligible patients were aged 30-99 years, and had at least one year of prior registration with the practice to ensure more accurate ascertainment of historical conditions.^30^

### Definition of long-term conditions

We examined 73 LTCs in women and 72 LTCs in men (two conditions only affect women, one condition only affects men, 71 conditions affect both), including all conditions recommended for inclusion in MLTC research where measurement was feasible in our routine data (Supplementary Table S1).^25^ These conditions were ascertained using lists of diagnostic codes (Read v2 for primary care data and ICD-10 in HES-APC and ONS data). Measurement was of the ‘lifetime history’ of each LTC, so in some cases the condition may not be active on the index date (for example, asthma and type 2 diabetes may remit and relapse). A small number of conditions were defined to be mutually exclusive (for example, we applied additional rules to define type 1 and type 2 diabetes as mutually exclusive when code lists flagged patients as having both),^31^ and where this was the case we removed associations from the model because the strong negative association observed is spurious. To facilitate visualisation, conditions were presented grouped into 14 categories defined by ICD-10 chapter.

### Inference of associations

Associations between LTCs are identified using Associations Beyond Chance (ABC), a Bayesian measure of non-random associations that controls for uncertainty.^14^ The ABC measure is conceptually similar to the commonly employed Observed-to-Expected ratio (OER, sometimes called lift),^17,37^ but more conservative in its estimate of strength of association where sample sizes are small and OER is unstable (for example, for rare conditions or smaller subgroups of participants).^14^ ABC values reflect the ratio of two conditions co-appearing due to common mechanisms (*f_ij_*) as compared to them co-appearing by chance: 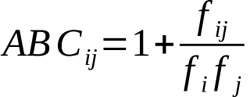. A value of *ABC_ij_=1* implies that co-appearance is likely due to chance, while *ABC_ij_>1* indicates that conditions co-occur more than expected by chance, or less than chance if *ABC_ij_<1*.

The values of *ABC_ij_*, *f_i_* and *f_j_* are inferred from LTC counts (*P_i_*), co-occurrence counts (*X_ij_*), and total number of individuals (*M*) within the group stratum. Inference is then achieved using a Bayesian approach:

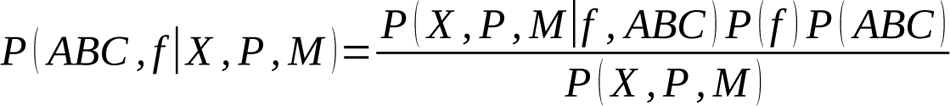

where the likelihood *P(X,P | f, ABC)* is defined by independent multinomials, the priors 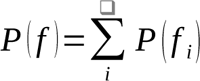 are defined as beta distributions with shared parameters α and β defined by respective exponential hyperpriors with *λ=0.01*, and the priors 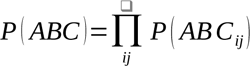 are defined by a log-normal distribution with *µ=0.25* and *σ=0.5*. This distribution was chosen to have its mode at *ABC_ij_=1* and 90^th^ percentile at 2.437, therefore rejecting the possibilities of strong non-random associations when evidence is scarce.

To avoid the costly computation of the evidence term *P(X, P, M)*, inference of the posterior was obtained via Hamiltonian Monte Carlo,^16^ using 2,000 warm-up iterations to allow the sampling to stabilize. We then sampled 1,000 values to characterize the distribution of inferred variables and excluded associations between pairs of conditions as non-significant if the 0.5^th^-99.5^th^ percentile interval of their *ABC* distribution included the value *ABC=1*. The reported values correspond to the mode of these distributions, which we estimate by the middle point of the shortest span of ten consecutive values in the 1,000 samples of each *ABC_ij_*.

### Initial analysis

All analyses were stratified by sex. For every possible pair of conditions, we calculated joint prevalence (numerator: number of individuals with both conditions present within the specified stratum; denominator: number of individuals in specified stratum) and the magnitude of statistically significant pairwise associations using ABC. In men and women separately, each of these three measures was calculated in all ages together (30-99 years), and then further stratified by age, grouped into six age bands: 30-39, 40-49, 50-59, 60-69, 70-79, 80-99 years.

### Network construction and community detection (condition clustering)

To further study how conditions cluster together in each sex and all ages together (30-99 years) versus age-sex stratified analyses, we built MLTC networks from ABC associations and applied a community-detection algorithm. In MLTC networks, each LTC is represented by a node and pairs of LTCs are connected if their ABC association is significant, with connection weights corresponding to the mode of the distribution of ABC values.

A problem in the analysis of association networks is that they usually have many weak links arising from unobserved factors, mediating effects, and errors in the data. Previous works using MLTC networks typically filter out connections whose weights are below a certain threshold or a specified percentile.^17,23^ This filtering approach is not ideal on association networks, where weight values are commonly correlated with condition prevalence, as this would risk biasing results against some conditions.^17,18^ Instead of threshold filtering, we applied *Noise-Corrected Backboning* (NCB), a Bayesian filtering approach that accounts for typical weight values of each node to only keep connections whose weights are confidently above typical values.^19^ The NCB filter has a parameter that controls for the tolerance to noise, and we have chosen a value of *δ=1.64*, corresponding approximately to a p-value of 0.05.^19^

Once we obtained the filtered network, we applied a modularity maximization algorithm to find communities, i.e., groups of conditions strongly associated in the network. Given an assignment of conditions to communities, with *c_i_=k* meaning that condition *i* is assigned to community *k*, the modularity *Q* of the assignment can be defined as

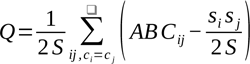

where the sum is over all pairs of nodes that are placed within the same community, *ABC_ij_* is the weight of the edge between conditions *i* and *j* after filtering, *s_i_* is the sum of association values of a condition 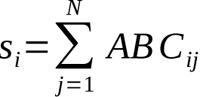, and *S* is the total sum of ABC values in the network: 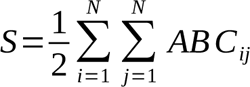. To find the condition clustering that yields the largest modularity value, ran the Leiden algorithm^20^ 10^5^ times per network (where we run the algorithm until convergence each time) and selected the result with the highest modularity value.

We measured similarity between different clustering assignments with the Adjusted Rand Index (ARI), where a value of zero reflects that clusters are no more similar than expected by random assignments, and a value of one corresponds to identical cluster assignments.^33^

### Ethics

Provision of anonymized data for research by the Clinical Practice Research Datalink (CPRD) is approved annually by the NHS Research Ethics Service, and study specific NREC review is not required provided the study has been approved by the CPRD Independent Scientific Advisory Committee (ISAC). This study was reviewed by ISAC and approved (protocol 21_000542).

## Results

### Study population

The study population consisted of 7,490,874 individuals aged 30-99 years. Women comprised 3,757,667 (50.2%) of participants, and 37.8% were aged 60-99 years, compared to 34.0% of the 3,733,207 male participants (supplementary Table S1). In all women aged 30-99 years, the most common conditions were hypertension (25.0%), anxiety (21.2%), depression (21.2%), asthma (16.4%), thyroid disease (10.4%), osteoporosis or major osteoporotic fracture (9.7%), chronic kidney disease (9.0%), and cataract (9.0%). In all men aged 30-99 years, the commonest conditions were hypertension (25.0%), asthma (13.4%), anxiety (12.1%), depression (12.0%), type 2 diabetes (9.0%), coronary heart disease (8.8%), benign prostatic hypertrophy (7.3%), and alcohol misuse (7.1%). Prevalence of individual LTCs varied considerably by age, most commonly increasing with age (for example hypertension, macular degeneration, dementia) but sometimes showing flatter or n-shaped patterns (for example depression, asthma) (supplementary Table S2).

### Pairwise associations and joint prevalence in the all-ages analysis (sex-stratified)

In all-ages, there was a median 65 (IQR 59 to 68) statistically significant pairwise associations per condition measured using ABC in men, and 65 (IQR 58 to 68) in women (Figure 1, supplementary Table S3). The median magnitude of association measured by ABC was 2.50 (IQR 1.65 to 3.74) in men, and 2.29 (IQR 1.49 to 3.47) in women, and the median joint prevalence of pairs of conditions was 0.031% (IQR 0.006 to 0.143) in men and 0.030% (IQR 0.006 to 0.143) in women.

Figure 2 shows the magnitude of pairwise association (measured using ABC) and joint prevalence in the whole population (30-99 years) stratified by sex, with conditions grouped by body system. In both men and women, there were multiple statistically significant positive associations for a large majority of diseases. For some conditions, there were strong associations with conditions in other body systems (for example, intellectual disability with epilepsy; dementia with Parkinson’s Disease, Addison’s Disease with type 1 diabetes), but the general pattern was of stronger positive associations between conditions in the same body system, particularly for mental health conditions, cardiovascular disease, respiratory disease, neurological disease, and chronic infections (HIV and tuberculosis) (supplementary Figure S1).

**Figure 1:**
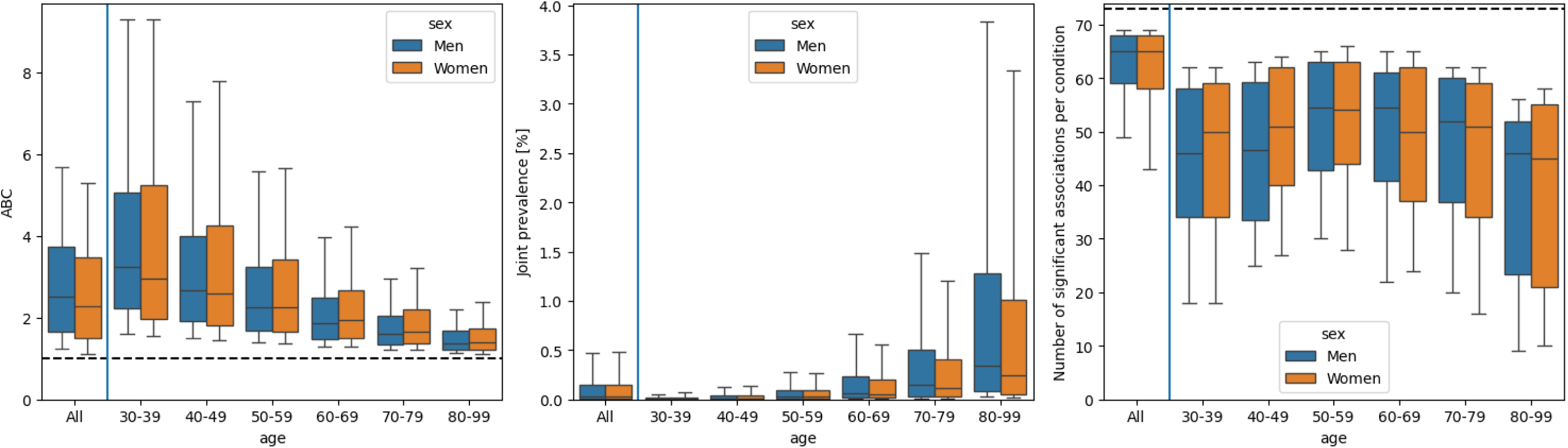
(a) ABC magnitude of significant associations, (b) joint prevalence, and (c) number of significant associations per condition, across age groups in men (blue) and women (orange). Boxplots include the median and quartiles, with whiskers extending to the 10th and 90th percentiles.

**Figure 2:**
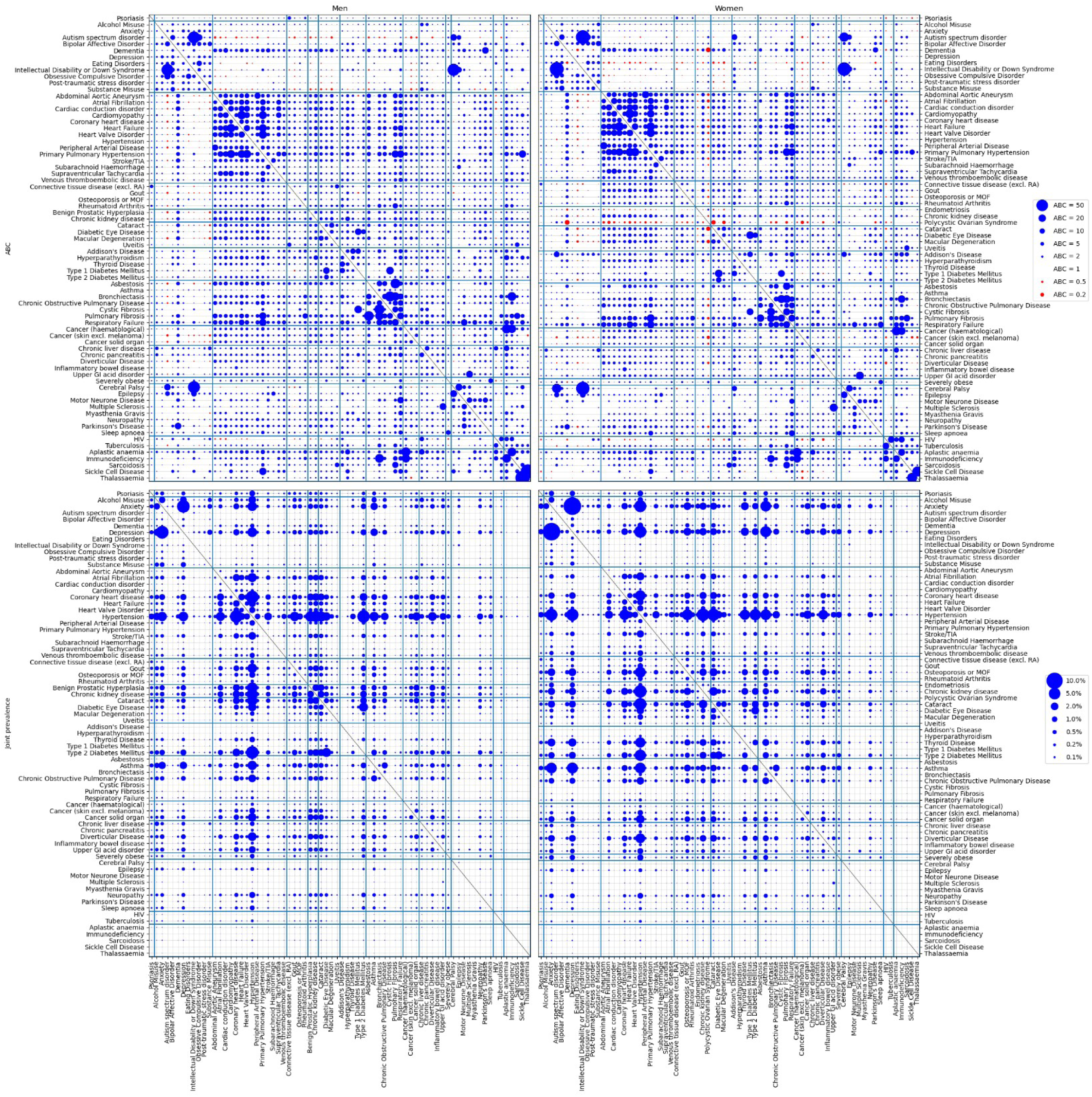
ABC associations (top) and joint prevalence (bottom) between all pairs of conditions for men (left column) and women (right column) of all ages (30-99 years). The bubble size (area) is proportional to the ABC magnitude of the association (after offsetting to 1 and inverting negative values) or joint prevalence. Blue bubbles represent statistically significant positive associations (greater than expected by chance), red bubbles represent statistically significant negative associations (less than expected by chance), and empty spots reflect no significant association.

A small number of conditions had statistically significant negative correlations with multiple other conditions from different body systems, notably several mental health conditions in both sexes (autism, eating disorders, intellectual disability, obsessive compulsive disorder, post-traumatic stress disorder, and substance misuse), two gynaecological disorders affecting pre-menopausal women (endometriosis and polycystic ovarian syndrome), and HIV (supplementary Figure S2)

As expected, joint prevalence was generally higher for pairs of common conditions, but there was again some evidence of higher joint prevalence for conditions in the same body system.

### Pairwise associations and joint prevalence stratified by age in addition to sex

In analyses further stratified by age, the number of statistically significant associations per condition increased with age, peaking in 50-59 year-olds in both sexes (median 54.5 in men, 54 in women), before reducing in the oldest age groups (46 in men, 45 in women in 80-99 year-olds) (Figure 1). Strength of association measured by ABC was highest in 30-39 year-olds (median ABC 3.23 in men, 2.95 in women) and declined progressively in older age groups (in 80-99 year-olds, 1.37 in men and 1.40 women). In contrast, joint prevalence of pairs of conditions was lowest in 30-39 year-olds (median [IQR] joint prevalence 0.005% [0.002-0.016] in men, 0.007% [0.002-0.024] in women) and progressively increased with age (in 80-99 year-olds, median [IQR] 0.341% [0.084-1.28] in men, 0.244% [0.053-1.010] in women).

Supplementary Figure S10 and S11 show the ten most prevalent pairs of conditions in each age group, with mental health conditions being prominent in the under-60s (either as pairs of mental health conditions, most commonly depression and anxiety, or as part of mental-physical multimorbidity) but physical condition pairs dominating in older age-groups.

Figure 3 shows all pairwise associations and joint prevalences for three age groups (30-39 years, 50-59 years and 80-99 years; all other age groups are shown in extended supplementary Figures S3-S9). Consistent with the data shown in Figure 1, in both men and women the magnitude of associations fell markedly with age while joint prevalence considerably increased (Figure 3). In all age groups, the strength of association was usually stronger within body system groups than between body system groups (supplementary Figure S1), particularly for mental health, circulatory, respiratory, eye, infectious and blood conditions, and particularly in younger age groups. Notably, there were fewer statistically significant negative associations in age-stratified analyses, and the conditions with consistently negative associations in the all-ages analysis had consistently positive associations in each age subgroup (supplementary Figure S2).

**Figure 3:**
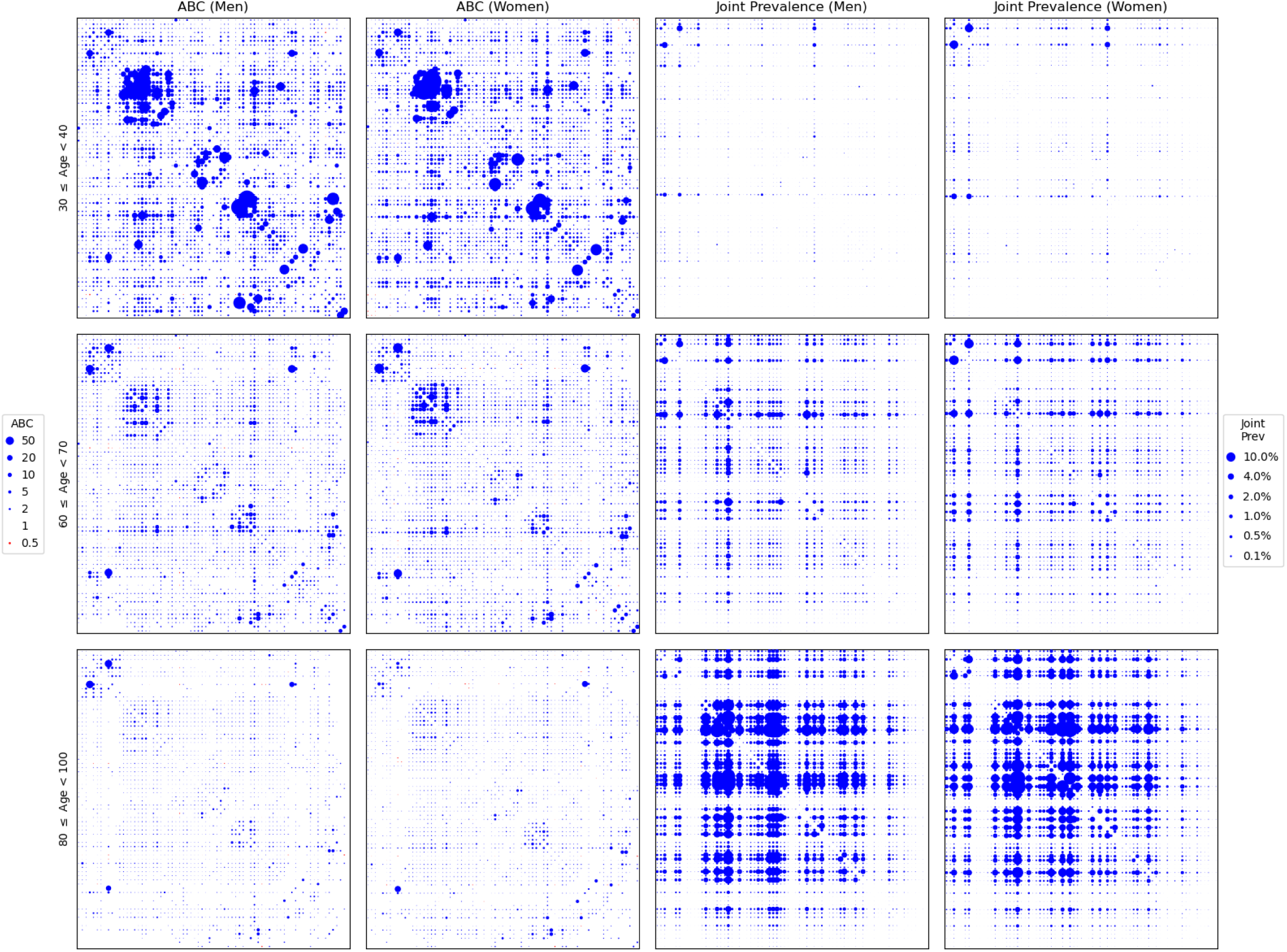
ABC associations (left) and joint prevalence (right) between all pairs of conditions for (a) men and (b) women in three age strata: 30-39yo (top), 60-69yo (middle), 80-99yo (bottom). Bubble sizes (area) are proportional to the ABC magnitude of the association (after offsetting to 1 and inverting negative values) or joint prevalence. Blue bubbles represent statistically significant positive associations (greater than expected by chance), red bubbles represent statistically significant negative associations (less than expected by chance), and empty spots reflect no significant association.

### Condition clustering in the all-ages analysis (sex-stratified)

In the all-ages analysis, there were eight clusters identified both in men and in women (Figures 4 and 6), although even after backboning there were still multiple significant connections between conditions in different clusters. Specifically, we observed mean 22.0 (SD 19.0) statistically significant connections within a cluster, and 23.9 (SD 17.8) connections to conditions outside the cluster, but strength of association within each cluster was stronger (median ABC 11.6 [1.8-23.7 10th-90th percentile]) than with conditions outside the cluster (median ABC 5.5 [1.6-10.9]). Two clusters were similar in both men and women (being dominated by cardiovascular and cancer/neurological conditions, although not including exactly the same conditions), and some others were similar to lesser degrees. For example, in men there were distinct mental health and alcohol/chronic liver disease clusters, whereas in women these conditions largely clustered together along with the two included gynaecological conditions. Furthermore, there were clusters observed in one sex which were not observed in the other, such as chronic kidney disease (CKD)/gout/hyperparathyroidism in women. In men, these three conditions were part of the endocrine/obesity cluster, whose other conditions were part of an overlapping but distinct respiratory/rheumatological/obesity cluster in women (Figure 6).

**Figure 4:**
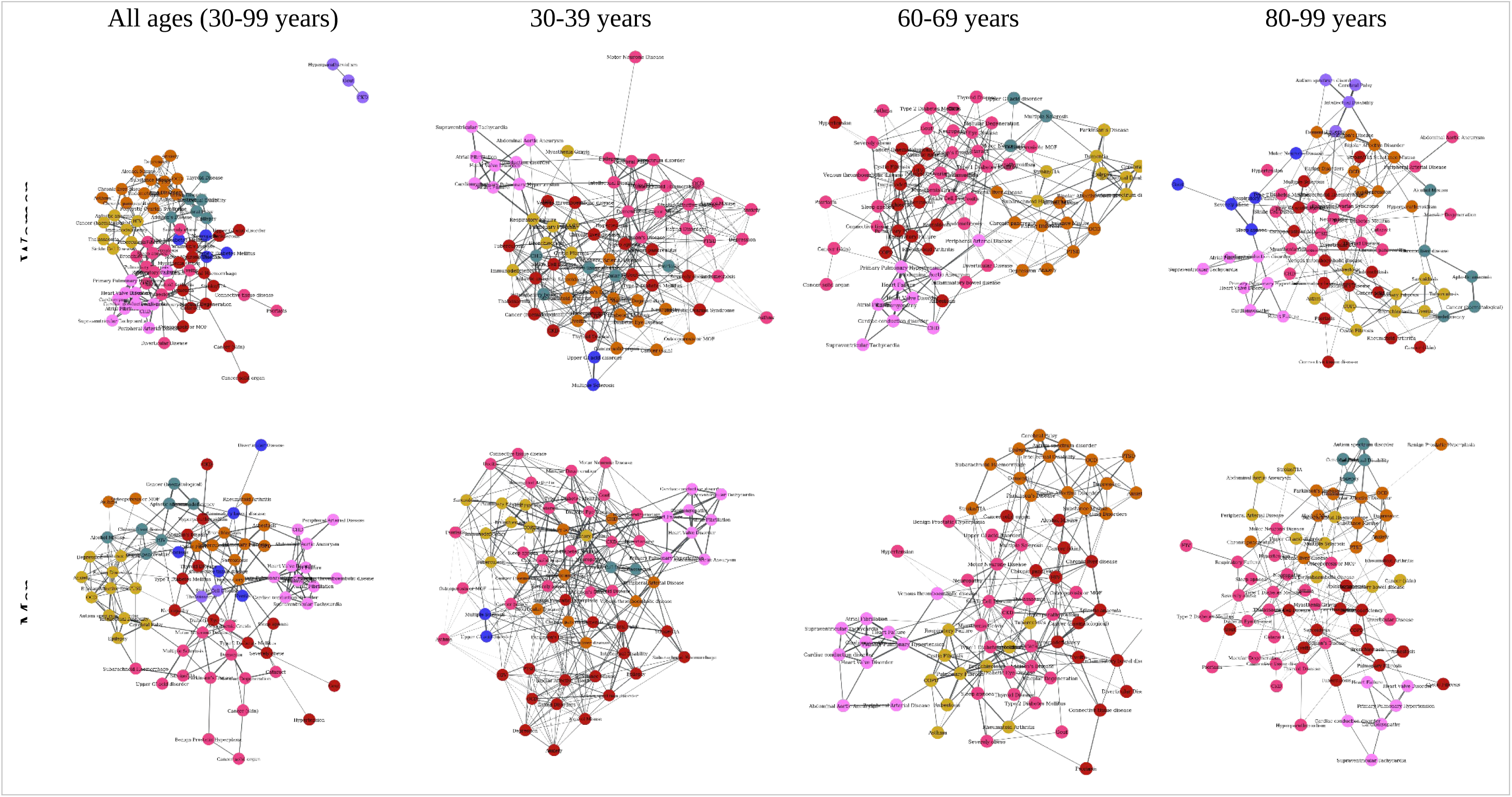
Networks of associations and condition clusters for men (top) and women (bottom) when looking at all ages (left-most) or selected age strata. The networks were built from significant ABC associations and applied a backboning noise-corrected filter. Condition clustering in each group was selected by the run with highest modularity of a modularity maximisation algorithm run 10^5^ times.

### Condition clustering stratified by age in addition to sex

After age-sex stratification, consistent with patterns of pairwise association, the networks were observed to be denser in younger people and less dense in older (Figure 4). In men, there were five clusters observed at ages 30-39 years and 60-69 years, and six at age 80-99 years, compared to seven, six and eight in women.

There was less consistency in how conditions clustered in different age groups in women than in men (the ARI cluster similarity between age groups was lower in women [0.27, 0.18, and 0.25] than in men [0.50, 0.40, and 0.34]), although in both sexes the same condition could appear in very different clusters at different ages and/or in the all-ages-grouped clustering (Figure 6). In both sexes, there was a ‘cardiovascular’ cluster consistently observed at all ages, although the conditions included in this cluster were not the same in every age group (Figure 6), and associations with conditions in other clusters varied by age group (Figure 5). Some other clusters were fairly consistently observed in different age groups but often with variation by sex and differences from the all-ages cluster. In men, there was a respiratory cluster observed in all-ages and in each age group, whereas in women respiratory conditions were reasonably consistently clustered in the age-stratified analysis but were part of a respiratory/rheumatology/obesity cluster in all-ages. Mental health conditions did cluster in both sexes at all-ages and in age strata, but the pattern of clustering was different in the age strata compared to all-ages. In both sexes, there were clusters observed in one age stratum which were not observed in others or in all-ages (for example, multiple sclerosis/upper GI acid disorder in both sexes in age 30-39 years).

**Figure 5:**
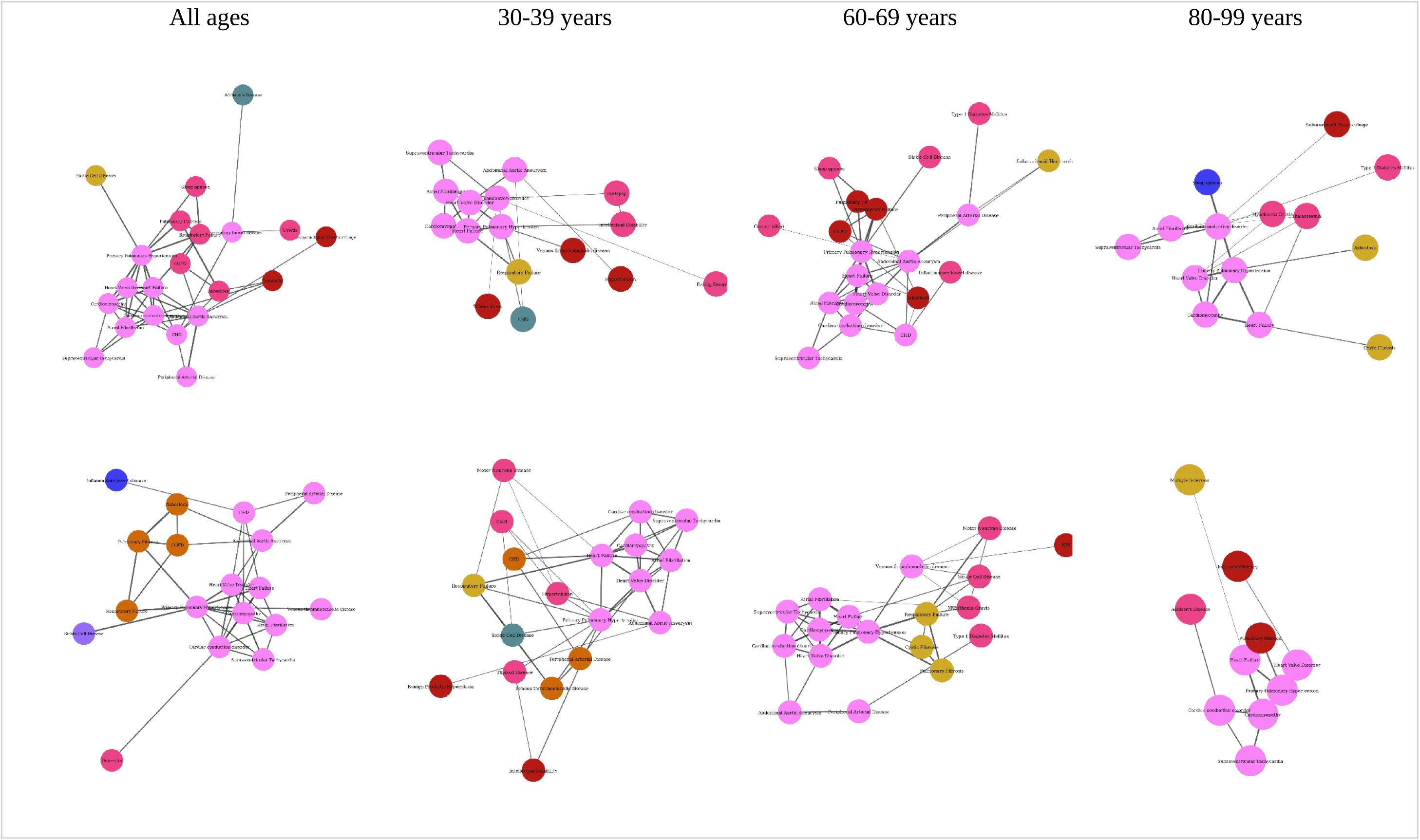
A close-up of the cardiovascular clusters (in pink) and their neighbouring conditions --- i.e. those with direct connections to any condition in the cluster --- from the networks shown in Figure 4.

**Figure 6.**
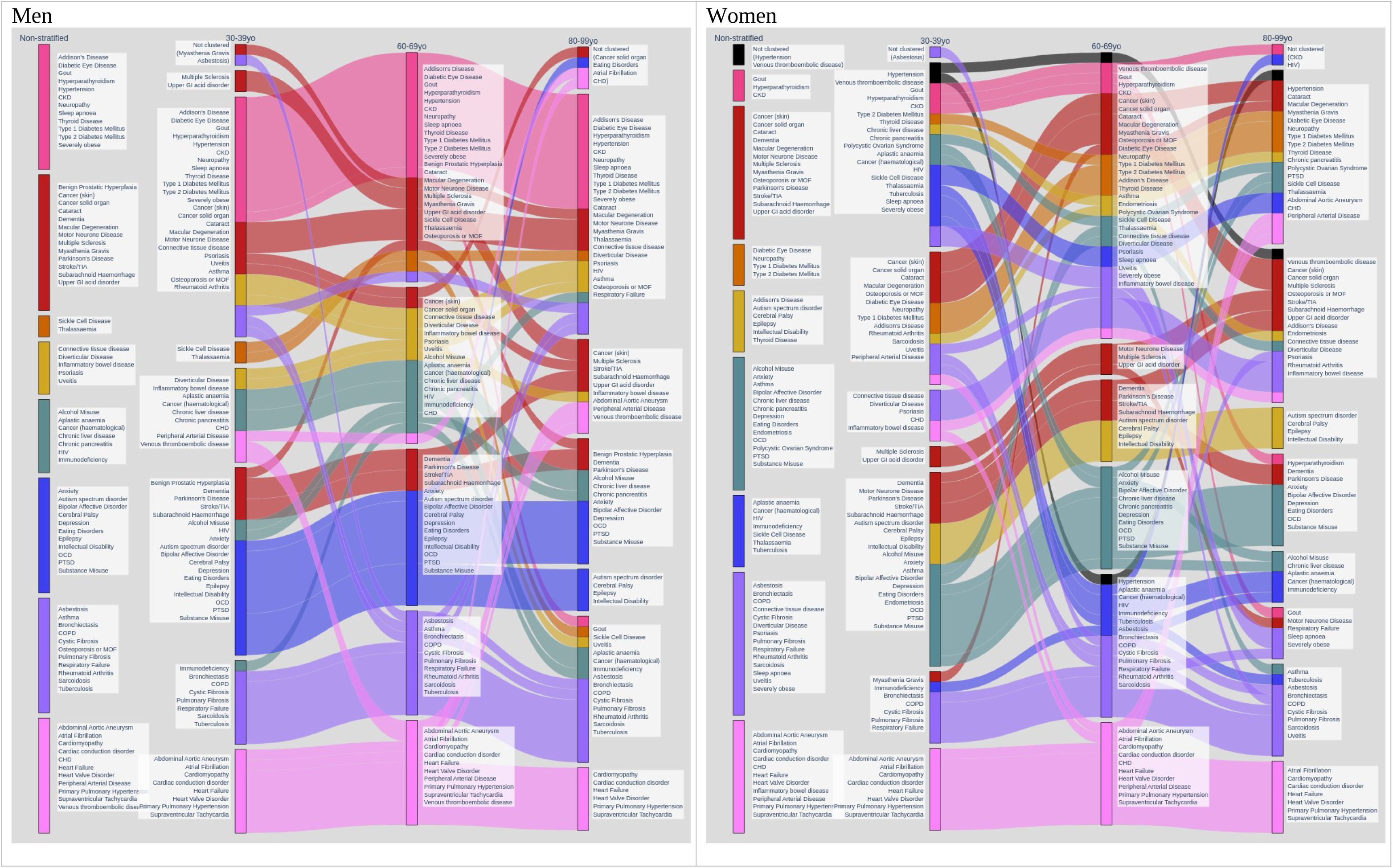
Cluster membership of conditions in all men and women (left-most column in both images), and for age groups 30-39 years, 60-79 years, and 80-99 years. Clusters are the same as shown in Figure 4.

## Discussion

This study finds that patterns of pairwise associations and joint prevalence are different in the whole population of men or women aged 30-99 years compared to age-stratified subgroups. Specifically, age-stratified analysis finds that the magnitude of pairwise association is systematically greater in younger age groups whereas joint prevalence is systematically larger in older age-groups. In the sex-stratified all-ages analyses, a number of statistically significant negative associations are observed which are not present in any of the age strata. Clusters of conditions identified in the all-ages analysis differ between men and women in multiple ways, and are similarly not stable across age-stratified groups, with greater heterogeneity between different age strata in women than men.

There is a growing body of research examining how LTCs cluster. These studies vary in terms of the conditions examined and how those conditions are measured in data, both of which make comparison across studies and reproducibility problematic.^5,6,21,32^ Our study shows that the comparison of cluster solutions across studies will further vary depending on choices made about the study population and stratification of analysis, which matters because there is also considerable variation between studies in terms of the age range of the datasets used, and whether or not clustering is age and/or sex stratified.^2,3,13,15,34,35^ A further issue not examined by this study is that the choice of a clustering method and measures of association will also affect the number and composition of the clusters identified, although cardiovascular and mental health clusters of various compositions are fairly consistently observed (as was the case in this study).^2,17,22^ Overall, this study highlights that interpretation and generalisability of condition clusters is highly uncertain.

Our study has several strengths. We were able to undertake analyses in a very large population dataset which is reasonably representative of the UK population,^26^ with ascertainment of conditions from both hospital and primary care.^21,32^ Furthermore, we evaluated a broad range of long-term conditions including those recommended for use in multimorbidity research by a recent consensus panel.^25^ While clustering approaches may simultaneously evaluate multiple features to create clusters, explainability is often problematic and clinical interpretation of cluster solutions is often unclear. Our approach of systematically examining understandable measures (prevalence and pairwise associations) in pairs of conditions (before undertaking one specific approach to clustering based on the ABC measure of association) provides greater interpretability of the differences observed between analyses undertaken in age-stratified compared with whole populations. Additionally, network visualisations of clusters, as shown in Figure 5, make it somewhat easier to ascertain the main associations that form (and explain) a cluster, as well as the neighbouring conditions that are not part of the cluster but closely associated with some of its constituent conditions.

A limitation of our study is that it uses routine data collected for clinical care, which is likely to be more subject to misdiagnosis and miscoding than prospectively collected research data, although CPRD has been shown to be valid for ascertainment of many conditions, and bespoke research datasets are typically much smaller and only collect data for fewer and less granular conditions. Second, although we included 74 conditions, these were sometimes ascertained at a relatively coarse level. As an example, we chose to combine multiple connective tissue disorders into a single category, and to define a mutually exclusive group with rheumatoid arthritis. Our rationale was that people with these conditions may acquire a number of diagnostic labels as their conditions evolve (for example, polymyalgic presentations of rheumatoid arthritis may initially be labelled as polymyalgia rheumatica), and clinical records may contain the whole ‘correct at the time’ history rather than being edited to only reflect final diagnosis. Granularity therefore carries risk of spurious association due to how conditions are diagnosed, labelled, and recorded in routine data. There is no definitive solution to this, but it is important that researchers are explicit in how and why they define conditions to include, as we do.^25,39^ Third, we defined conditions as ‘lifetime history’ rather than currently active, although for aetiological research our approach is likely appropriate. Fourth, we have only applied a single clustering method and cluster solutions are likely more persuasive if consistently observed by different methods, although in this paper we are only using clustering for demonstration purposes, and therefore discourage over-interpreting the clustering results. Fifth, some of the differences between whole-population and age-stratified clustering may reflect that age-stratified datasets are necessarily smaller and therefore have less power to detect associations.

Finally, although we compare clusters observed in different age groups, it is important to recognize that this cross-sectional study is not examining how conditions change with age in individuals, since the patients in each age group come from different generational cohorts, and cohort or period effects will potentially confound any implied age effects.^24^ Longitudinal studies have greater ability to observe how conditions cluster within individuals over time. For example, Dervic et al used longitudinal hospital admission data to construct temporal networks, and identified 1260 distinct trajectories observed over a mean of 23 years, including some sets of trajectories in younger people which diverge as they age.^23^ However, longitudinal clustering poses its own problems, including that the true sequence of conditions is rarely known for certain, particularly where conditions are derived from codes recorded in hospital discharge data, which is usually very non-granular in terms of date of diagnosis, as well as having incomplete ascertainment of many conditions, notably mental health.^21,32^

Our findings demonstrate the importance for multimorbidity researchers of considering the age range and structure of the populations in which analyses using clustering methods are undertaken, and careful consideration of the intended application of their findings. The importance of age stratification before applying clustering reflects the variation in patterns of disease with both age and time (i.e. period/cohort as well as age effects). Conditions vary in the ages when they are most commonly incident, but this will be affected by changes in risk factors which will have variably applied to different cohorts. For example, the age-specific experience of smoking and obesity has changed considerably over time (current 30-39 year-olds are less likely to smoke and more likely to be obese than current 60-69 year-olds were 30 years ago).

Similarly, life expectancy for people with conditions like intellectual disability, type 1 diabetes, and childhood cancer has improved considerably, as has survival from acute life-threatening conditions like myocardial infarction and common cancers in middle and older age. Furthermore, participants are, by definition, the subset of survivors who have not experienced a terminal condition like severe heart failure or untreatable cancer. Patterns of association in different age groups are therefore expected to be different, which will be compounded by changes in data recording. For example, we observed multiple negative associations in analyses of whole populations for some conditions (a range of mental health conditions commoner in younger people and pre-menopausal gynaecological conditions) but not in any of the individual age strata. This is likely due to changes in diagnostic practice and data recording over time. Autism spectrum disorder for example shows steep declines in recorded prevalence with increasing age (supplementary Table S2), reflecting changing diagnostic criteria and potentially no *electronic* record of diagnosis in the distant past, even if the diagnosis was made. Researchers therefore need to carefully consider which conditions to include in analysis, and how changes in diagnostic practice and electronic recording of data may influence observed associations and clusters.

Researchers need to understand how their choices about which populations and conditions to examine may influence the results of whatever clustering method they apply. The lack of consistency of some clusters between whole populations and across age strata in our study may indicate that, at least for studies intending to promote aetiological understanding, whole population clustering is likely to be misleading. At a minimum, researchers should explore how clustering varies by age and sex. Similarly, we believe that the sensitivity of clustering to data and methodological choices, and the heterogeneity of clusters in different population strata limits any immediate clinical application without further methodologically focused research and validation of cluster solutions.^13^ Currently, we believe that understanding joint prevalence and simple description of common patterns has more immediate implications for health services planning and individual care than strength of association, which is why clinical interest in multimorbidity often focuses on older people (where joint prevalence is highest) and selected groups of younger people where there are clear clinical implications (such as early onset cardiometabolic disease). Clustering based on strength of association does have promise for exploring aetiology, but most existing research seeking to cluster MLTC does not provide explainable solutions that easily inform understanding.

## Conclusion

Pairwise measures of association and joint prevalence for LTCs vary between age strata and whole populations of women and men, resulting in clusters of LTCs that are not consistently reproducible across populations. Alongside explicit justification of the LTCs examined,^25^ researchers should carefully consider their choice of population and whether stratification by key variables such as age and sex is required before undertaking clustering analyses of MLTCs.

## Data Availability

The data controllers do not allow sharing of raw data by the research team, but any researcher can access the same raw data subject to data controller approval (https://www.cprd.com/). Code lists for defining the conditions examined can be found online at Zenodo.^38^

## Acknowledgements

The study was funded by the National Institute for Health Research (NIHR) Artificial Intelligence and Multimorbidity: Clustering in Individuals, Space and Clinical Context (NIHR202639). The funder had no role in conduct of the study, interpretation, or the decision to submit for publication. The views expressed are those of the authors and not necessarily those of the NIHR, the Department of Health and Social Care. This study is based in part on data from the Clinical Practice Research Datalink obtained under licence from the UK Medicines and Healthcare products Regulatory Agency. The data is provided by patients and collected by the NHS as part of their care and support. The interpretation and conclusions contained in this study are those of the author/s alone The authors thank Milos Nikolic for assisting with the database management.

## Author contributions

Funding for the AIM-CISC programme was obtained by BG, VR, NZ and JF, and the analysis reported conceived by GRM, VR, NL, JF and BG. Initial data management was led by LDF, JP and BG, and analysis by GRM, VR, JF, BG and NL. All authors contributed to interpretation and drafting of the manuscript, and approved the final version.

## Competing interests

There are no competing interests.

## Notes

### Competing Interest Statement

The authors have declared no competing interest.

