## Supplementary Materials for "Pitfalls in understanding how multiple long-term conditions cluster: whole population and age-stratified associations in 7,490,874 people in England"

Supplementary Figure S2: ABC magnitude of significant NEGATIVE associations, (b) their joint prevalence, and (c) number of significant NEGATIVE associations per condition, across age groups and in men (blue) and women (orange). Boxplots include the median and quartiles, with whiskers extending to the 10th and 90th percentiles...9

Supplementary Table S1: Study population demography and prevalence of long-term conditions

| | **Men**<br>**No. (%) of men**<br>**(N=3733207)** | **Women**<br>**No. (%) of women**<br>**(N=3757667)** |
|---|---|---|
| Age-group | | |
| 30-39 | 841765 (22.55%) | 806156 (21.45%) |
| 40-49 | 809411 (21.68%) | 758652 (20.19%) |
| 50-59 | 813714 (21.80%) | 773995 (20.60%) |
| 60-69 | 588217 (15.76%) | 588123 (15.65%) |
| 70-79 | 438586 (11.75%) | 482340 (12.84%) |
| 80-99 | 241514 (6.47%) | 348401 (9.27%) |
| Hypertension | 931457 (24.95%) | 938170 (24.97%) |
| Anxiety | 451758 (12.10%) | 798601 (21.25%) |
| Depression | 446508 (11.96%) | 797283 (21.22%) |
| Asthma | 498276 (13.35%) | 616713 (16.41%) |
| Type 2 Diabetes Mellitus | 334481 (8.96%) | 269652 (7.18%) |
| Chronic kidney disease | 243048 (6.51%) | 338388 (9.01%) |
| Cataract | 237256 (6.36%) | 336635 (8.96%) |
| Coronary heart disease | 327962 (8.78%) | 223098 (5.94%) |
| Osteoporosis or MOF | 171122 (4.58%) | 366055 (9.74%) |
| Thyroid Disease | 95505 (2.56%) | 390720 (10.40%) |
| Diverticular Disease | 196350 (5.26%) | 243980 (6.49%) |
| Cancer (solid organ) | 183587 (4.92%) | 238767 (6.35%) |
| Alcohol Misuse | 263402 (7.06%) | 152643 (4.06%) |
| Psoriasis | 156495 (4.19%) | 161194 (4.29%) |
| Atrial Fibrillation | 171725 (4.60%) | 131367 (3.50%) |
| Chronic Obstructive Pulmonary Disease | 150803 (4.04%) | 147335 (3.92%) |
| Gout | 233861 (6.26%) | 61967 (1.65%) |
| Benign Prostatic Hyperplasia | 274172 (7.34%) | 0 |
| Diabetic Eye Disease | 143512 (3.84%) | 109917 (2.93%) |
| Upper GI acid disorder | 136996 (3.67%) | 102828 (2.74%) |
| Stroke/TIA | 121545 (3.26%) | 111979 (2.98%) |
| Neuropathy | 110565 (2.96%) | 111247 (2.96%) |
| Venous thromboembolic disease | 80428 (2.15%) | 102804 (2.74%) |
| Cancer (skin excl. melanoma) | 95023 (2.55%) | 85053 (2.26%) |
| Severely obese | 55206 (1.48%) | 120656 (3.21%) |
| Heart Failure | 97487 (2.61%) | 76727 (2.04%) |
| Heart Valve Disorder | 77354 (2.07%) | 76404 (2.03%) |
| Epilepsy | 69367 (1.86%) | 71896 (1.91%) |
| Chronic liver disease | 74343 (1.99%) | 60917 (1.62%) |
| Substance Misuse | 90951 (2.44%) | 42777 (1.14%) |
| Endometriosis | 0 | 130971 (3.49%) |
| Connective tissue disease (excl. RA) | 50356 (1.35%) | 74081 (1.97%) |

| | | |
|---|---|---|
| Macular Degeneration | 44058 (1.18%) | 70142 (1.87%) |
| Rheumatoid Arthritis | 31919 (0.86%) | 74577 (1.98%) |
| Dementia | 40660 (1.09%) | 64871 (1.73%) |
| Peripheral Arterial Disease | 59999 (1.61%) | 35957 (0.96%) |
| Sleep apnoea | 66217 (1.77%) | 26990 (0.72%) |
| Inflammatory bowel disease | 42898 (1.15%) | 47074 (1.25%) |
| Uveitis | 36475 (0.98%) | 38011 (1.01%) |
| Supraventricular Tachycardia | 25241 (0.68%) | 36384 (0.97%) |
| Tuberculosis | 28636 (0.77%) | 30985 (0.82%) |
| Polycystic Ovarian Syndrome | 0 | 58348 (1.55%) |
| Cancer (haematological) | 31652 (0.85%) | 24490 (0.65%) |
| Chronic pancreatitis | 27059 (0.72%) | 28920 (0.77%) |
| Bronchiectasis | 23727 (0.64%) | 31328 (0.83%) |
| Intellectual Disability or Down Syndrome | 28777 (0.77%) | 22462 (0.60%) |
| Respiratory Failure | 22374 (0.60%) | 23266 (0.62%) |
| Post-traumatic stress disorder | 21240 (0.57%) | 19350 (0.51%) |
| Bipolar Affective Disorder | 15903 (0.43%) | 24400 (0.65%) |
| Obsessive Compulsive Disorder | 14775 (0.40%) | 21041 (0.56%) |
| Cardiomyopathy | 20817 (0.56%) | 11166 (0.30%) |
| Type 1 Diabetes Mellitus | 17673 (0.47%) | 12897 (0.34%) |
| Abdominal Aortic Aneurysm | 22481 (0.60%) | 5635 (0.15%) |
| Eating Disorders | 1489 (0.04%) | 26341 (0.70%) |
| Parkinson's Disease | 15495 (0.42%) | 11297 (0.30%) |
| Cardiac conduction disorder | 15778 (0.42%) | 10688 (0.28%) |
| Hyperparathyroidism | 6609 (0.18%) | 18268 (0.49%) |
| Multiple Sclerosis | 6890 (0.18%) | 17334 (0.46%) |
| Sarcoidosis | 10564 (0.28%) | 10509 (0.28%) |
| Subarachnoid Haemorrhage | 8324 (0.22%) | 10185 (0.27%) |
| Pulmonary Fibrosis | 9502 (0.25%) | 7764 (0.21%) |
| Autism spectrum disorder | 8899 (0.24%) | 3304 (0.09%) |
| Primary Pulmonary Hypertension | 3892 (0.10%) | 5641 (0.15%) |
| Thalassaemia | 2882 (0.08%) | 5997 (0.16%) |
| Cerebral Palsy | 4571 (0.12%) | 4003 (0.11%) |
| Asbestosis | 7554 (0.20%) | 460 (0.01%) |
| Aplastic anaemia | 4078 (0.11%) | 3878 (0.10%) |
| HIV | 3957 (0.11%) | 2273 (0.06%) |
| Immunodeficiency | 1931 (0.05%) | 2410 (0.06%) |
| Sickle Cell Disease | 1370 (0.04%) | 2437 (0.06%) |
| Addison’s Disease | 1339 (0.04%) | 2416 (0.06%) |
| Myasthenia Gravis | 1843 (0.05%) | 1861 (0.05%) |
| Motor Neurone Disease | 1121 (0.03%) | 777 (0.02%) |
| Cystic Fibrosis | 443 (0.01%) | 572 (0.02%) |

Conditions listed in descending order of prevalence in combined population of men and women

MOF – major osteoporotic fracture; TIA – transient ischaemic attack; RA – rheumatoid arthritis; HIV – Human Immunodeficiency Virus. Conditions with zero cases for men or for women are specific to the other sex.

Supplementary Table S2: Prevalence of conditions by age-group in all participants

| | All participants (both women and men) (N=7490874) | 30-39 % with LTC N= 1647921 | 40-49 % with LTC N= 1568063 | 50-59 % with LTC N= 1587709 | 60-69 % with LTC N= 1176340 | 70-79 % with LTC N= 920926 | 80-99 % with LTC N= 589915 |
|---|---|---|---|---|---|---|---|
| Hypertension | 1869627 (24.96%) | 1.95 | 7.71 | 19.84 | 37.70 | 56.66 | 73.94 |
| Anxiety | 1250359 (16.69%) | 15.00 | 16.93 | 17.87 | 17.59 | 16.82 | 15.61 |
| Depression | 1243791 (16.60%) | 12.81 | 16.91 | 19.25 | 18.88 | 16.63 | 14.69 |
| Asthma | 1114989 (14.88%) | 16.78 | 14.92 | 14.15 | 13.74 | 14.23 | 14.77 |
| Type 2 Diabetes Mellitus | 604133 (8.06%) | 0.82 | 3.09 | 7.07 | 12.78 | 17.32 | 20.35 |
| Chronic kidney disease | 581436 (7.76%) | 1.39 | 1.71 | 2.89 | 6.70 | 17.52 | 41.63 |
| Cataract | 573891 (7.66%) | 0.29 | 0.63 | 1.90 | 6.57 | 18.66 | 47.46 |
| Coronary heart disease | 551060 (7.36%) | 0.57 | 1.37 | 4.15 | 9.91 | 18.04 | 29.09 |
| Osteoporosis or MOF | 537177 (7.17%) | 3.28 | 3.09 | 4.32 | 7.85 | 13.27 | 25.70 |
| Thyroid Disease | 486225 (6.49%) | 2.42 | 4.03 | 5.89 | 8.48 | 11.12 | 14.82 |
| Diverticular Disease | 440330 (5.88%) | 0.23 | 0.92 | 3.76 | 7.82 | 14.89 | 22.58 |
| Cancer (solid organ) | 422354 (5.64%) | 0.64 | 1.54 | 3.70 | 7.80 | 14.10 | 18.19 |
| Alcohol Misuse | 416045 (5.55%) | 4.55 | 5.52 | 6.53 | 6.76 | 5.60 | 3.31 |
| Psoriasis | 317689 (4.24%) | 3.04 | 3.79 | 4.42 | 5.06 | 5.40 | 4.84 |
| Atrial Fibrillation | 303092 (4.05%) | 0.19 | 0.48 | 1.32 | 3.86 | 10.02 | 22.67 |
| Chronic Obstructive Pulmonary Disease | 298138 (3.98%) | 0.20 | 0.75 | 2.47 | 6.14 | 10.45 | 12.77 |
| Gout | 295828 (3.95%) | 0.62 | 1.72 | 3.38 | 5.75 | 8.17 | 10.56 |
| Benign Prostatic Hyperplasia | 274172 (3.66%) | 0.04 | 0.23 | 1.31 | 4.84 | 10.84 | 15.66 |
| Diabetic Eye Disease | 253429 (3.38%) | 0.45 | 1.20 | 2.71 | 5.17 | 7.41 | 9.32 |
| Upper GI acid disorder | 239824 (3.20%) | 0.58 | 1.32 | 2.63 | 4.57 | 6.63 | 8.98 |
| Stroke/TIA | 233524 (3.12%) | 0.27 | 0.63 | 1.56 | 3.56 | 7.27 | 14.55 |
| Neuropathy | 221812 (2.96%) | 0.80 | 1.68 | 2.98 | 4.35 | 5.32 | 5.93 |
| Venous thromboembolic disease | 183232 (2.45%) | 0.73 | 1.30 | 1.91 | 2.94 | 4.66 | 7.29 |
| Cancer (skin excl. melanoma) | 180076 (2.40%) | 0.11 | 0.40 | 1.05 | 2.54 | 5.92 | 12.02 |
| Severely obese | 175862 (2.35%) | 1.60 | 2.32 | 3.05 | 3.12 | 2.33 | 1.10 |
| Heart Failure | 174214 (2.33%) | 0.12 | 0.29 | 0.87 | 2.31 | 5.27 | 13.25 |
| Heart Valve Disorder | 153758 (2.05%) | 0.23 | 0.40 | 0.83 | 1.98 | 4.69 | 10.85 |
| Epilepsy | 141263 (1.89%) | 1.55 | 1.77 | 1.98 | 2.05 | 2.10 | 2.20 |
| Chronic liver disease | 135260 (1.81%) | 0.84 | 1.61 | 2.27 | 2.60 | 2.23 | 1.52 |
| Substance Misuse | 133728 (1.79%) | 2.72 | 2.76 | 1.76 | 0.90 | 0.51 | 0.41 |
| Endometriosis | 130971 (1.75%) | 1.28 | 2.42 | 2.60 | 1.66 | 0.87 | 0.50 |
| Connective tissue disease (any excl. RA) | 124437 (1.66%) | 0.38 | 0.64 | 1.04 | 1.81 | 3.48 | 6.48 |
| Macular Degeneration | 114200 (1.52%) | 0.05 | 0.12 | 0.32 | 1.00 | 3.21 | 11.03 |
| Rheumatoid Arthritis | 106496 (1.42%) | 0.28 | 0.59 | 1.16 | 2.06 | 3.03 | 3.74 |
| Dementia | 105531 (1.41%) | 0.02 | 0.05 | 0.14 | 0.55 | 2.53 | 12.26 |
| Peripheral Arterial Disease | 95956 (1.28%) | 0.05 | 0.15 | 0.57 | 1.70 | 3.40 | 5.52 |
| Sleep apnoea | 93207 (1.24%) | 0.40 | 0.93 | 1.62 | 2.10 | 1.79 | 0.89 |
| Inflammatory bowel disease | 89972 (1.20%) | 0.80 | 1.00 | 1.20 | 1.46 | 1.70 | 1.58 |
| Uveitis | 74486 (0.99%) | 0.43 | 0.72 | 1.02 | 1.30 | 1.53 | 1.81 |
| Supraventricular Tachycardia | 61625 (0.82%) | 0.31 | 0.43 | 0.65 | 1.02 | 1.55 | 2.24 |
| Tuberculosis | 59621 (0.80%) | 0.46 | 0.59 | 0.58 | 0.85 | 1.24 | 2.04 |

| | | | | | | | |
|---|---|---|---|---|---|---|---|
| Polycystic Ovarian Syndrome | 58348 (0.78%) | 1.98 | 1.17 | 0.38 | 0.08 | 0.03 | 0.01 |
| Cancer (haematological) | 56142 (0.75%) | 0.19 | 0.30 | 0.53 | 1.00 | 1.65 | 2.19 |
| Chronic pancreatitis | 55979 (0.75%) | 0.31 | 0.50 | 0.70 | 0.92 | 1.22 | 1.64 |
| Bronchiectasis | 55055 (0.73%) | 0.09 | 0.18 | 0.37 | 0.96 | 2.00 | 2.57 |
| Intellectual Disability or Down Syndrome | 51239 (0.68%) | 0.84 | 0.71 | 0.78 | 0.66 | 0.46 | 0.30 |
| Respiratory Failure | 45640 (0.61%) | 0.12 | 0.21 | 0.40 | 0.80 | 1.35 | 2.06 |
| Post-traumatic stress disorder | 40590 (0.54%) | 0.61 | 0.75 | 0.67 | 0.46 | 0.23 | 0.10 |
| Bipolar Affective Disorder | 40303 (0.54%) | 0.45 | 0.58 | 0.62 | 0.59 | 0.52 | 0.38 |
| Obsessive Compulsive Disorder | 35816 (0.48%) | 0.65 | 0.59 | 0.48 | 0.38 | 0.29 | 0.19 |
| Cardiomyopathy | 31983 (0.43%) | 0.09 | 0.17 | 0.36 | 0.63 | 0.94 | 1.03 |
| Type 1 Diabetes Mellitus | 30570 (0.41%) | 0.44 | 0.48 | 0.48 | 0.39 | 0.27 | 0.18 |
| Abdominal Aortic Aneurysm | 28116 (0.38%) | 0.01 | 0.02 | 0.05 | 0.41 | 1.05 | 2.11 |
| Eating Disorders | 27830 (0.37%) | 0.56 | 0.56 | 0.38 | 0.21 | 0.09 | 0.04 |
| Parkinson’s Disease | 26792 (0.36%) | 0.01 | 0.02 | 0.09 | 0.35 | 1.04 | 1.90 |
| Cardiac conduction disorder | 26466 (0.35%) | 0.03 | 0.04 | 0.09 | 0.24 | 0.75 | 2.45 |
| Hyperparathyroidism | 24877 (0.33%) | 0.05 | 0.11 | 0.23 | 0.43 | 0.72 | 1.21 |
| Multiple Sclerosis | 24224 (0.32%) | 0.15 | 0.30 | 0.42 | 0.47 | 0.40 | 0.21 |
| Sarcoidosis | 21073 (0.28%) | 0.08 | 0.20 | 0.35 | 0.42 | 0.43 | 0.35 |
| Subarachnoid Haemorrhage | 18509 (0.25%) | 0.06 | 0.13 | 0.25 | 0.38 | 0.46 | 0.47 |
| Pulmonary Fibrosis | 17266 (0.23%) | 0.02 | 0.03 | 0.09 | 0.24 | 0.60 | 1.13 |
| Autism spectrum disorder | 12203 (0.16%) | 0.33 | 0.18 | 0.15 | 0.09 | 0.04 | 0.02 |
| Primary pulmonary hypertension | 9533 (0.13%) | 0.02 | 0.03 | 0.06 | 0.12 | 0.27 | 0.66 |
| Thalassaemia | 8879 (0.12%) | 0.14 | 0.14 | 0.11 | 0.10 | 0.10 | 0.10 |
| Cerebral Palsy | 8574 (0.11%) | 0.16 | 0.13 | 0.12 | 0.10 | 0.07 | 0.04 |
| Asbestosis | 8014 (0.11%) | 0.00 | 0.00 | 0.01 | 0.09 | 0.39 | 0.55 |
| Aplastic anaemia | 7956 (0.11%) | 0.04 | 0.06 | 0.08 | 0.14 | 0.18 | 0.28 |
| HIV | 6230 (0.08%) | 0.05 | 0.13 | 0.14 | 0.07 | 0.02 | 0.01 |
| Immunodeficiency | 4341 (0.06%) | 0.04 | 0.04 | 0.05 | 0.07 | 0.11 | 0.10 |
| Sickle Cell Disease | 3807 (0.05%) | 0.07 | 0.07 | 0.06 | 0.03 | 0.02 | 0.02 |
| Addison's Disease | 3755 (0.05%) | 0.03 | 0.04 | 0.05 | 0.06 | 0.07 | 0.07 |
| Myasthenia Gravis | 3704 (0.05%) | 0.01 | 0.02 | 0.04 | 0.06 | 0.11 | 0.14 |
| Motor Neurone Disease | 1898 (0.03%) | 0.01 | 0.01 | 0.02 | 0.04 | 0.06 | 0.06 |
| Cystic Fibrosis | 1015 (0.01%) | 0.02 | 0.01 | 0.01 | 0.01 | 0.01 | 0.01 |

Conditions listed in descending order of prevalence in combined population of men and women
MOF - major osteoporotic fracture; TIA – transient ischaemic attack; RA – rheumatoid arthritis; HIV – Human Immunodeficiency Virus

Supplementary Table S3: Data for Figure 1

| Measure | Agegroup (years) | Sex | | Minimum | 10th percentile | Lower quartile | Median | Upper quartile | 90th percentile | Maximum |
|---|---|---|---|---|---|---|---|---|---|---|
| Magnitude of association (ABC) | All (30-99) | Men | | 0.30 | 1.25 | 1.65 | 2.50 | 3.74 | 5.69 | 82.93 |
| | All (30-99) | Women | | 0.10 | 1.11 | 1.49 | 2.29 | 3.47 | 5.29 | 80.56 |
| | 30-39 | Men | | 0.40 | 1.61 | 2.23 | 3.23 | 5.06 | 9.28 | 285.20 |
| | 40-49 | Men | | 0.60 | 1.49 | 1.90 | 2.67 | 3.99 | 7.28 | 140.87 |
| | 50-59 | Men | | 0.46 | 1.39 | 1.67 | 2.25 | 3.25 | 5.58 | 60.04 |
| | 60-69 | Men | | 0.49 | 1.30 | 1.47 | 1.86 | 2.49 | 3.98 | 66.33 |
| | 70-79 | Men | | 0.42 | 1.20 | 1.34 | 1.60 | 2.04 | 2.96 | 99.52 |
| | 80-99 | Men | | 0.42 | 1.13 | 1.22 | 1.37 | 1.69 | 2.21 | 56.67 |
| | 30-39 | Women | | 0.43 | 1.54 | 1.96 | 2.95 | 5.23 | 9.29 | 391.13 |
| | 40-49 | Women | | 0.44 | 1.45 | 1.82 | 2.60 | 4.26 | 7.79 | 205.60 |
| | 50-59 | Women | | 0.49 | 1.37 | 1.64 | 2.25 | 3.41 | 5.66 | 105.32 |
| | 60-69 | Women | | 0.57 | 1.30 | 1.50 | 1.94 | 2.67 | 4.25 | 92.44 |
| | 70-79 | Women | | 0.50 | 1.22 | 1.36 | 1.64 | 2.19 | 3.21 | 86.91 |
| | 80-99 | Women | | 0.45 | 1.12 | 1.22 | 1.40 | 1.73 | 2.38 | 37.61 |
| Joint prevalence (%) | All | Men | | 0.000 | 0.002 | 0.006 | 0.031 | 0.143 | 0.472 | 6.725 |
| | All | Women | | 0.000 | 0.002 | 0.006 | 0.030 | 0.143 | 0.481 | 12.761 |
| | 30-39 | Men | | 0.000 | 0.001 | 0.002 | 0.005 | 0.016 | 0.054 | 5.525 |
| | 40-49 | Men | | 0.001 | 0.002 | 0.004 | 0.012 | 0.041 | 0.122 | 7.199 |
| | 50-59 | Men | | 0.001 | 0.003 | 0.007 | 0.025 | 0.098 | 0.276 | 7.790 |
| | 60-69 | Men | | 0.001 | 0.006 | 0.016 | 0.062 | 0.233 | 0.667 | 10.982 |
| | 70-79 | Men | | 0.001 | 0.011 | 0.034 | 0.143 | 0.502 | 1.488 | 18.983 |
| | 80-99 | Men | | 0.002 | 0.028 | 0.084 | 0.341 | 1.277 | 3.833 | 33.858 |
| | 30-39 | Women | | 0.000 | 0.001 | 0.002 | 0.007 | 0.024 | 0.077 | 10.954 |
| | 40-49 | Women | | 0.001 | 0.002 | 0.004 | 0.013 | 0.044 | 0.136 | 13.544 |
| | 50-59 | Women | | 0.001 | 0.003 | 0.007 | 0.025 | 0.089 | 0.266 | 14.814 |
| | 60-69 | Women | | 0.001 | 0.005 | 0.014 | 0.050 | 0.197 | 0.561 | 13.853 |
| | 70-79 | Women | | 0.001 | 0.009 | 0.028 | 0.111 | 0.412 | 1.211 | 14.329 |
| | 80-99 | Women | | 0.001 | 0.019 | 0.053 | 0.244 | 1.010 | 3.350 | 40.141 |
| Number of statistically significant associations | All | Men | | 25 | 47.2 | 59 | 65 | 68 | 69.9 | 71 |
| | All | Women | | 26 | 41.4 | 58 | 65 | 68 | 69.8 | 72 |
| | 30-39 | Men | | 0 | 18 | 34 | 46 | 58 | 62 | 66 |
| | 40-49 | Men | | 0 | 23.2 | 33.5 | 46.5 | 59.25 | 63 | 68 |
| | 50-59 | Men | | 7 | 30 | 42.75 | 54.5 | 63 | 65 | 68 |
| | 60-69 | Men | | 8 | 18.4 | 40.75 | 54.5 | 61 | 65.9 | 68 |
| | 70-79 | Men | | 1 | 19.1 | 36.75 | 52 | 60 | 62.9 | 65 |
| | 80-99 | Men | | 0 | 8.1 | 23.25 | 46 | 52 | 56 | 64 |
| | 30-39 | Women | | 0 | 16.4 | 34 | 50 | 59 | 62.8 | 68 |
| | 40-49 | Women | | 0 | 21.4 | 40 | 51 | 62 | 64.8 | 69 |

| | | | | | | | | | | |
|---|---|---|---|---|---|---|---|---|---|---|
| | 50-59 | Women | | 0 | 26.4 | 44 | 54 | 63 | 66 | 69 |
| | 60-69 | Women | | 5 | 24 | 37 | 50 | 62 | 65 | 67 |
| | 70-79 | Women | | 3 | 11.2 | 34 | 51 | 59 | 62 | 66 |
| | 80-99 | Women | | 0 | 6.8 | 21 | 45 | 55 | 58.8 | 62 |

Supplementary Figure S1: Median magnitude of significance associations within the conditions of a body system (solid lines) or to conditions in other body systems (dashed lines) in men (blue) and women (orange). Plotted points on the left-most side of the plot represent the values without age stratification.

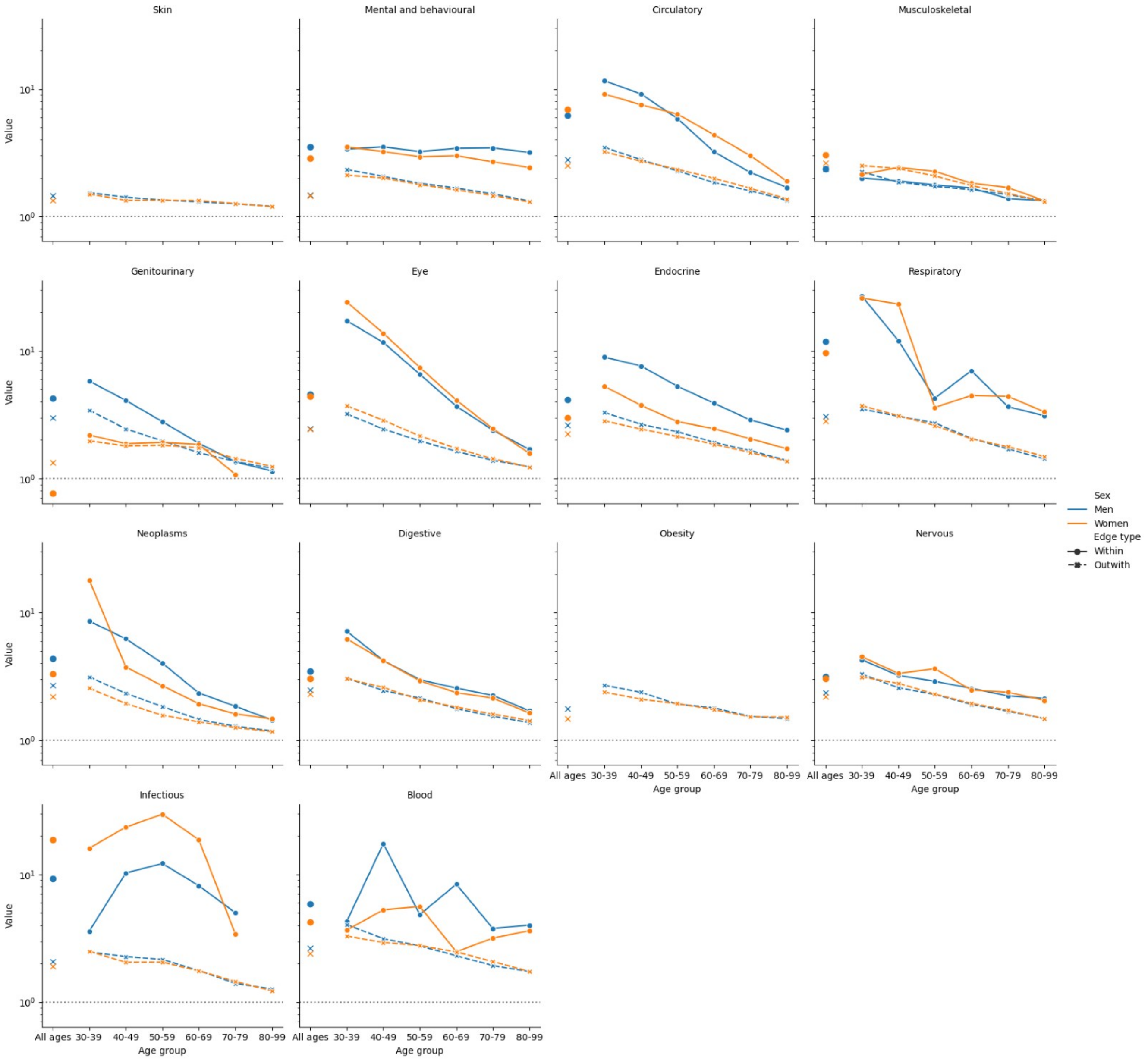

Supplementary Figure S2: ABC magnitude of significant NEGATIVE associations, (b) their joint prevalence, and (c) number of significant NEGATIVE associations per condition, across age groups and in men (blue) and women (orange). Boxplots include the median and quartiles, with whiskers extending to the 10th and 90th percentiles.

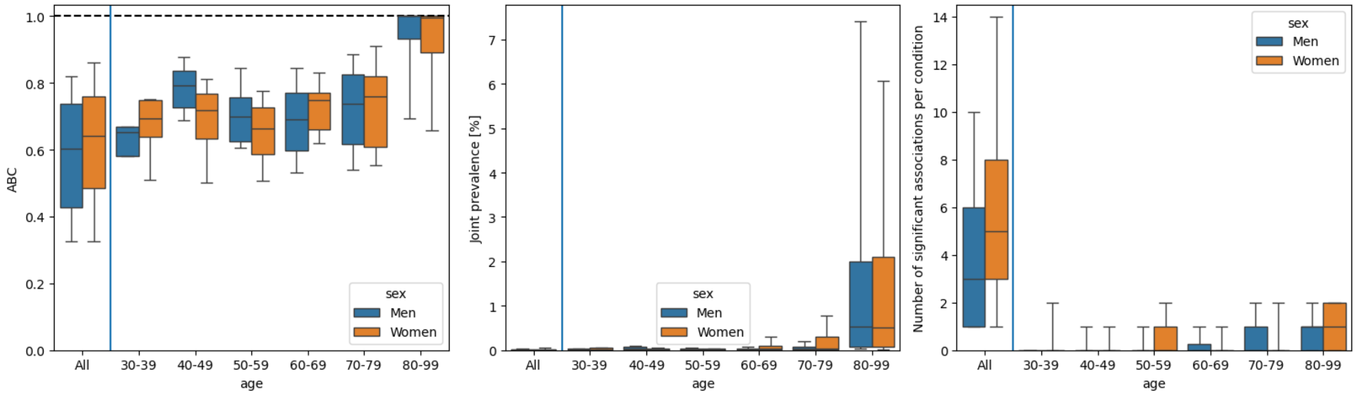

Supplementary figure S3: Strength of association (ABC) and joint prevalence in all age groups.

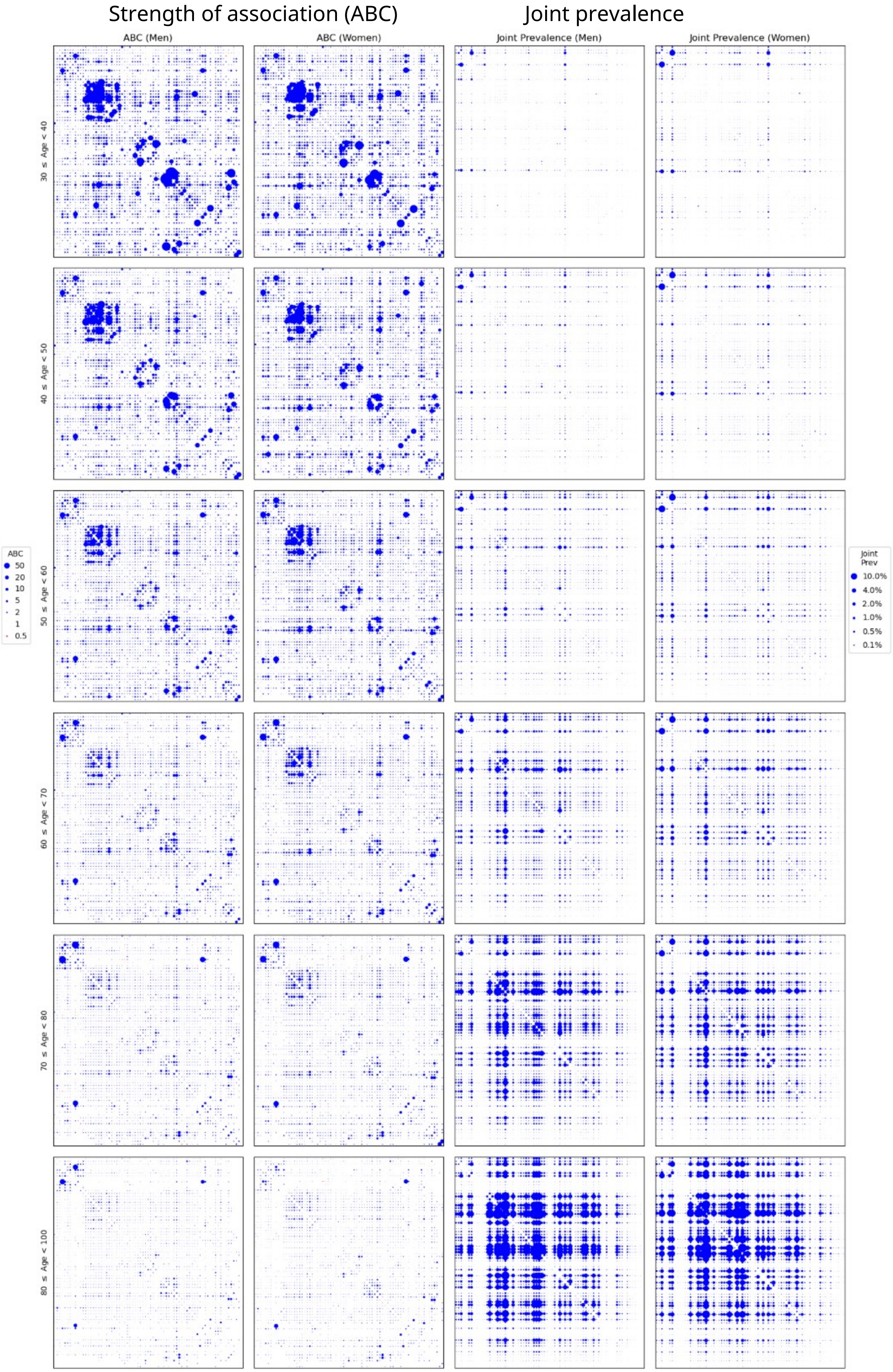

Supplementary figure S4: Strength of association (ABC) and joint prevalence in 30-39 year olds.

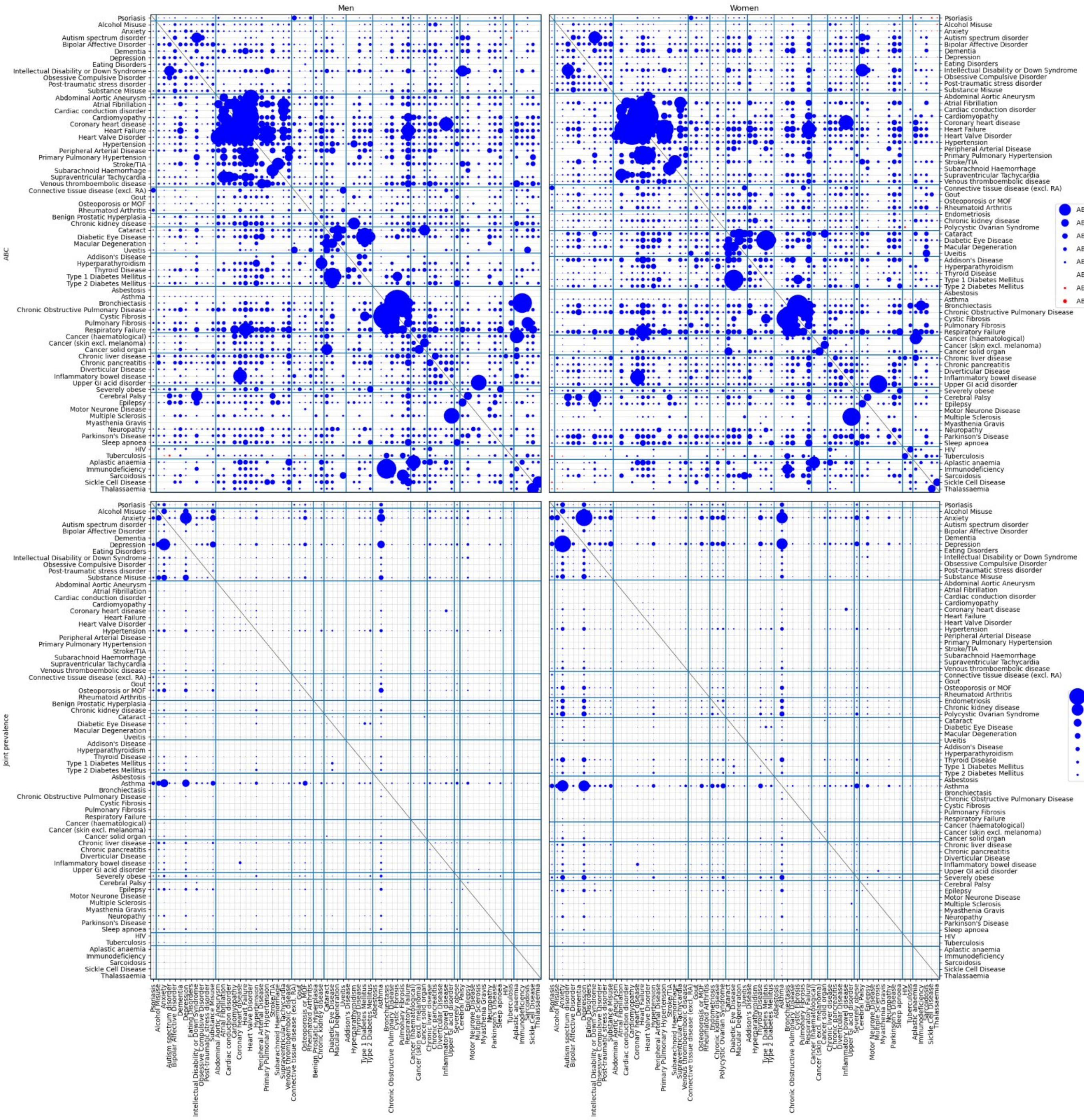

Supplementary figure S5: Strength of association (ABC) and joint prevalence in 40-49 year olds.

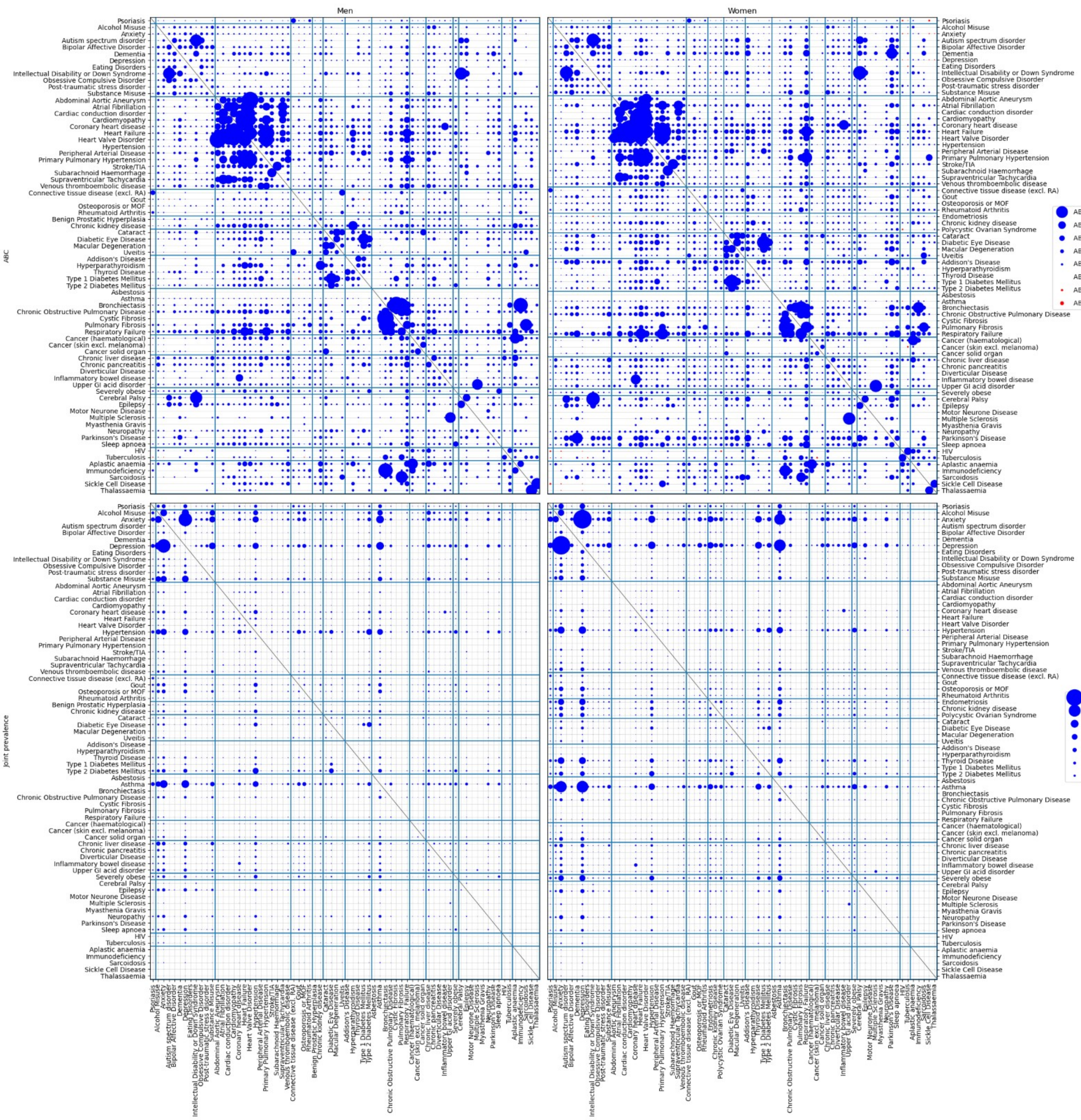

Supplementary figure S6: Strength of association (ABC) and joint prevalence in 50-59 year olds.

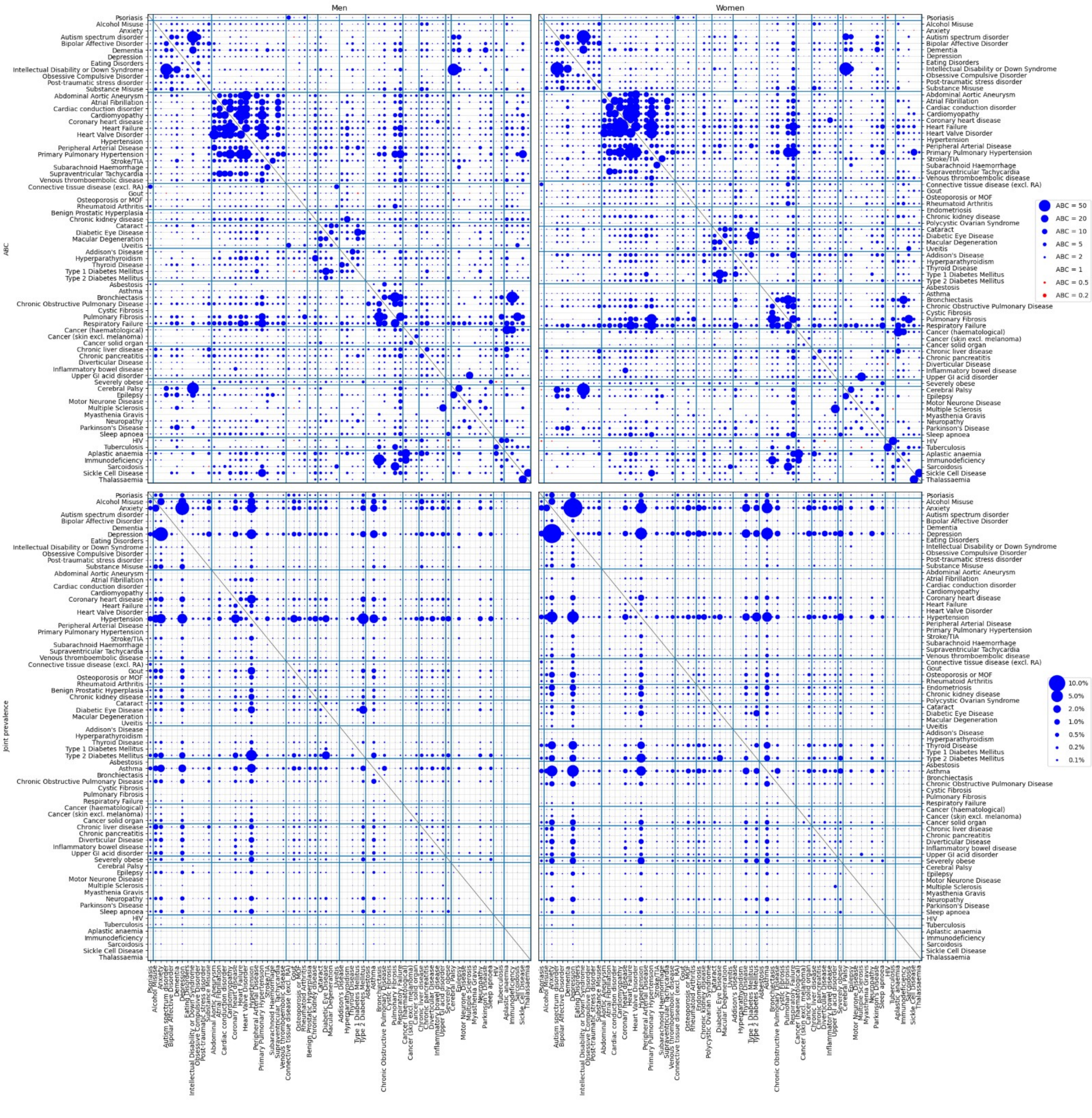

Supplementary figure S7: Strength of association (ABC) and joint prevalence in 60-69 year olds.

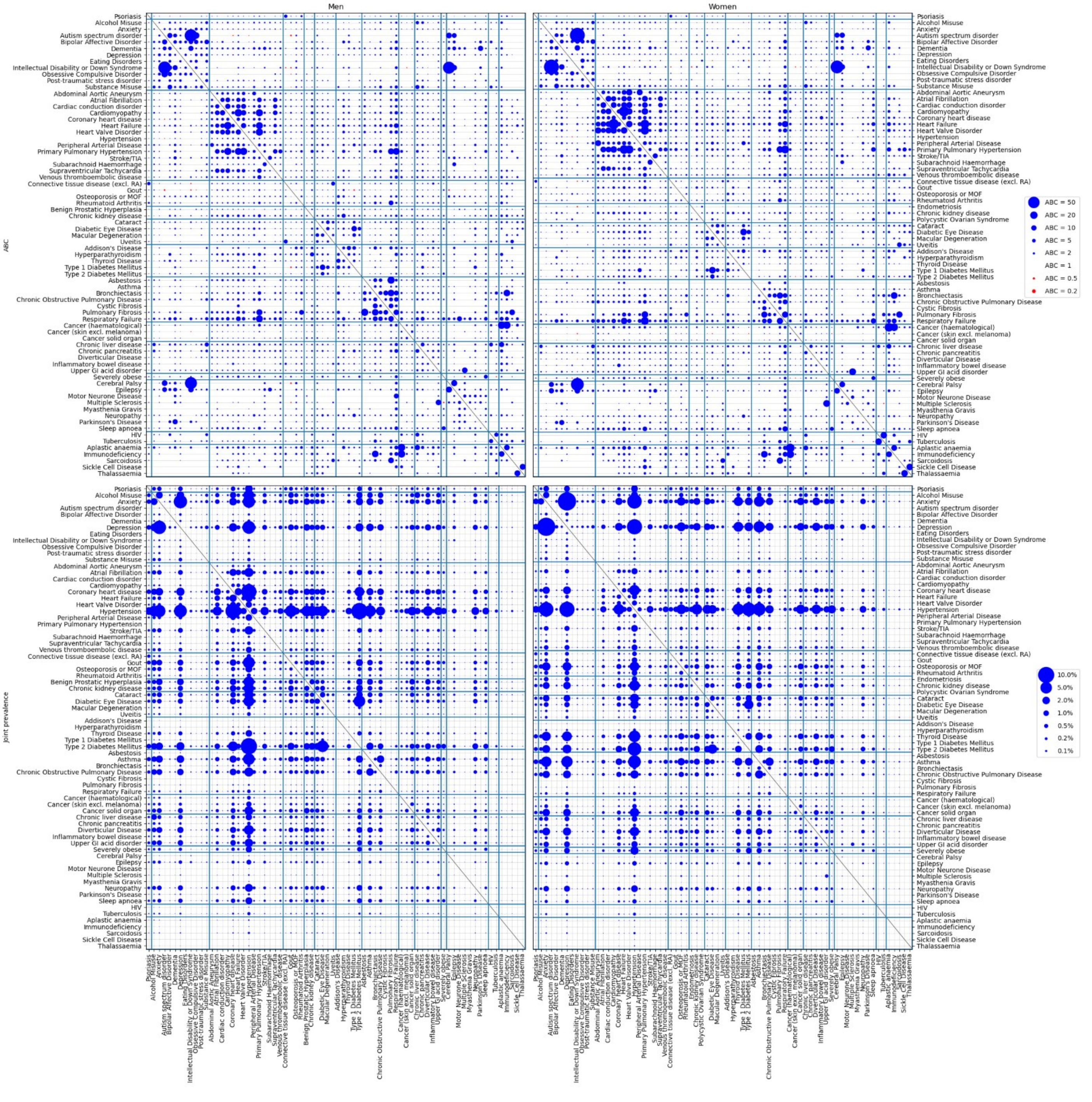

Supplementary figure S8: Strength of association (ABC) and joint prevalence in 70-79 year olds.

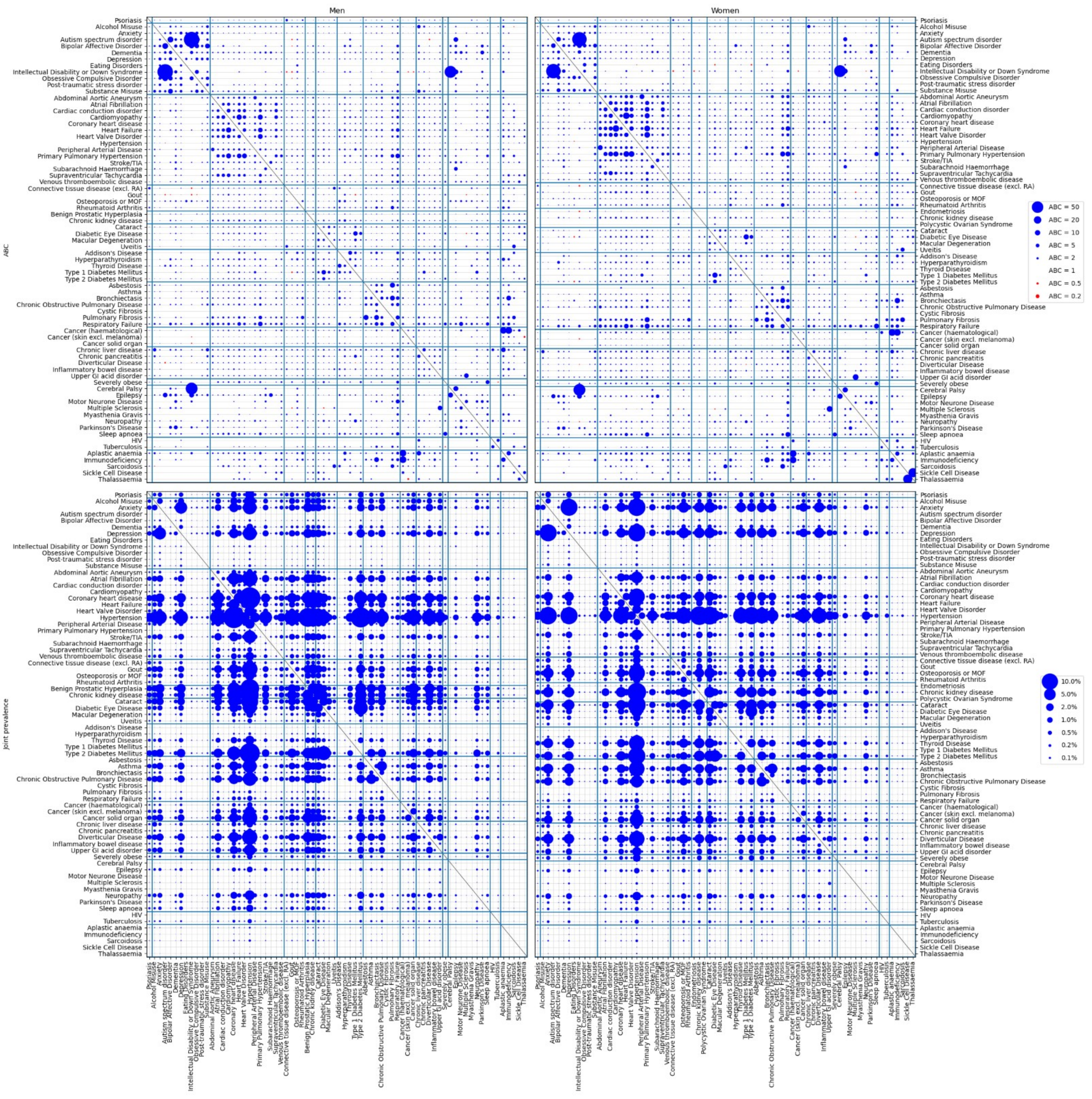

Supplementary figure S9: Strength of association (ABC) and joint prevalence in 80-99 year olds.

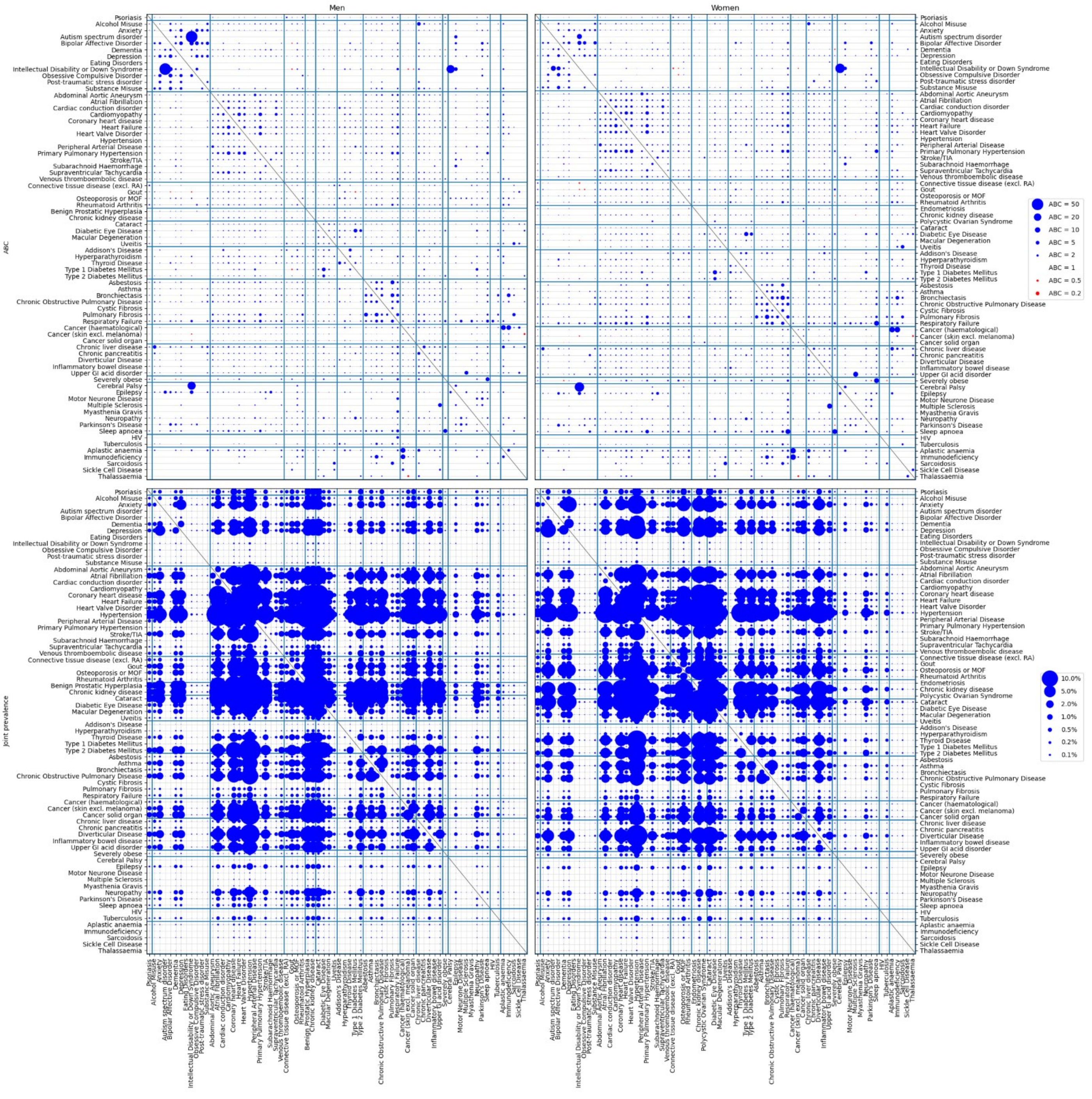

Supplementary Figure S10: Most frequent condition pairs in each age-group in men. Labels include the values of their joint prevalence and bubble size (area) are proportional to the strength of their association (ABC value).

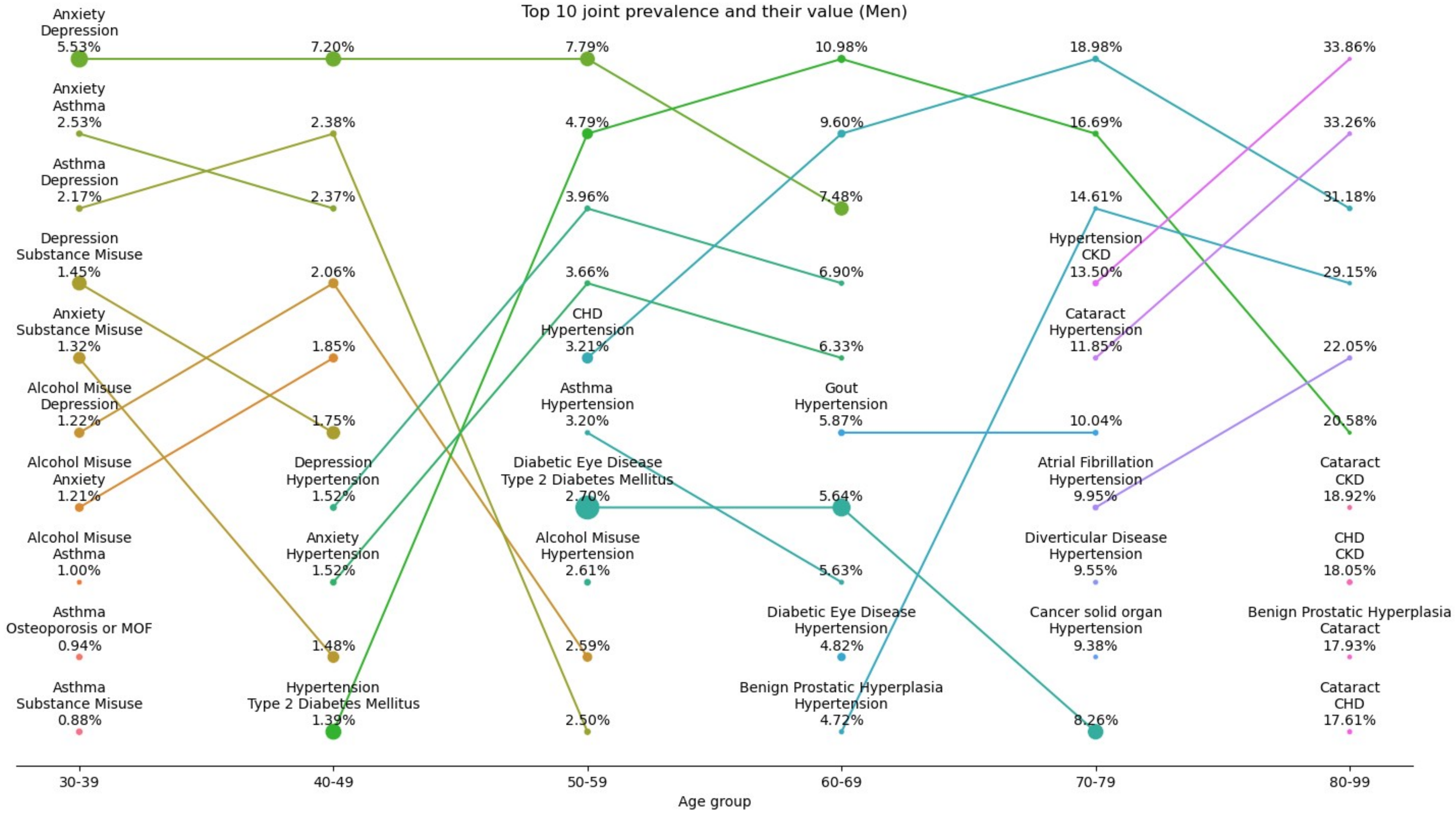

Supplementary Figure S11: Most frequent condition pairs in each age-group in women. Labels include the values of their joint prevalence and bubble size (area) are proportional to the strength of their association (ABC value).

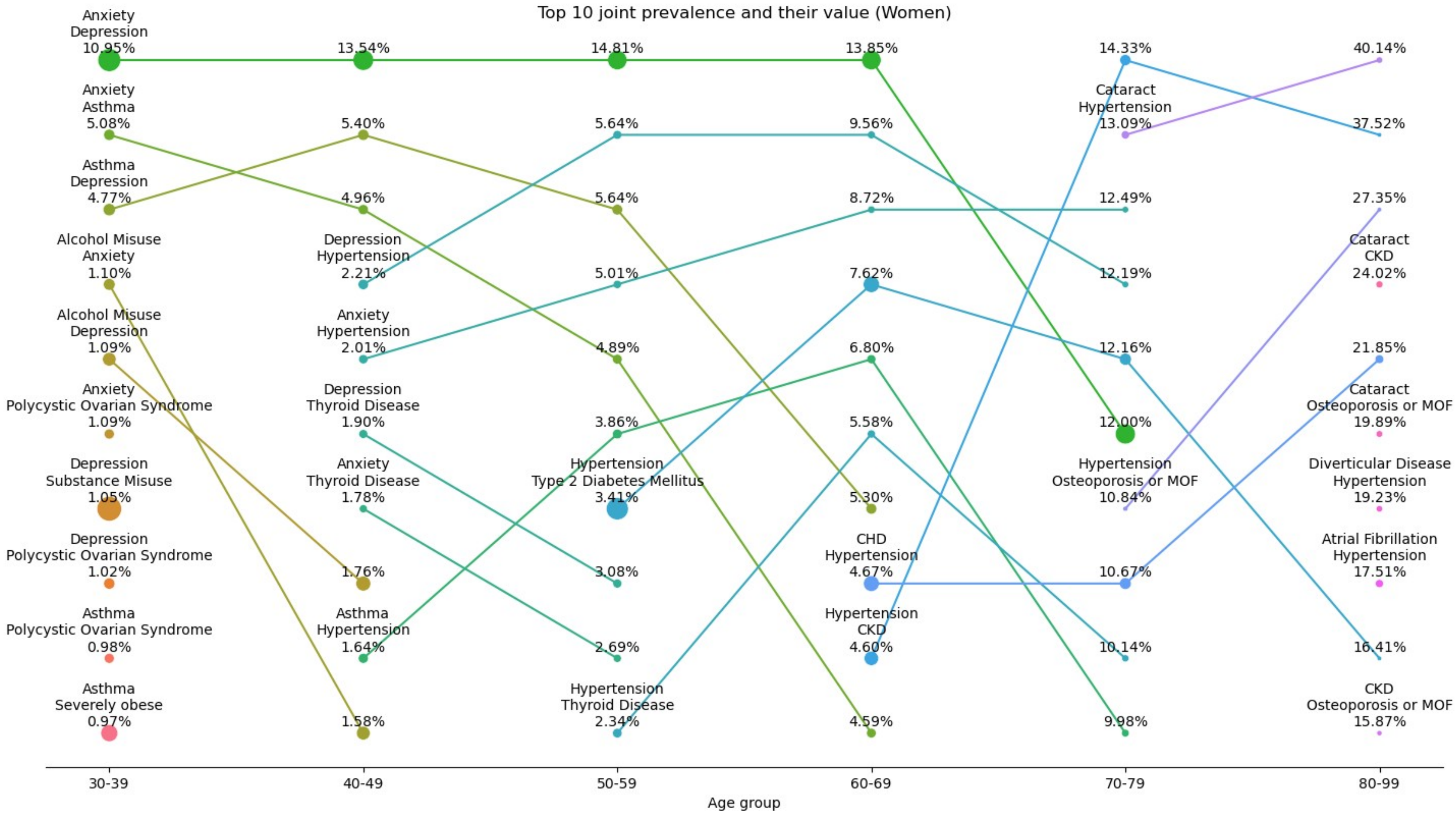
